# A geospatial approach to understanding transmission patterns of onchocerciasis in Ghana

**DOI:** 10.64898/2026.07.27.26359079

**Authors:** Millicent Opoku, David C. Deane, Himal Shrestha, Kwadwo K. Frempong, Sellase Pi-Bansa, Joseph Harold Nyarko Osei, Joseph Kwadwo Larbi Opare, Ernest Mensah, Odame D. Aseidu, Dziedzom K. de Souza, Daniel A. Boakye, Shannon M. Hedtke

## Abstract

**Background:** Onchocerciasis (river blindness), caused by the filarial nematode *Onchocerca volvulus* and transmitted by *Simulium* blackflies, remains a public health challenge in sub-Saharan Africa. Following the transition from vector control to large-scale ivermectin distribution, microfilariae (mf) prevalence has declined substantially across Ghana. The World Health Organization targets onchocerciasis elimination, defined as sustained mf prevalence below 1%. This requires identifying areas of persistent or resurgent transmission to guide targeted mapping surveys and interventions.

**Methods:** A Bayesian spatiotemporal, multilevel, generalized additive model with a zero-inflated beta-binomial outcome was fitted to mf prevalence data from 1353 surveys across 671 villages and environmental and socio-economic predictors. Temporal prevalence trends for ecozones within operational transmission zones (OTZ) were modelled using a 2D spatial smooth to account for residual spatial autocorrelation. Temporal autocorrelation in prevalence was accounted for using a random effect for village nested within ecozone.

**Results:** Runoff probability and surface wetness were positively associated with prevalence, consistent with *Simulium* breeding in fast-flowing water. Despite high initial mf prevalence, by 2015 the median prevalence across all OTZs in Ghana was estimated to be between 0.43 and 1.97%. The probability that prevalence was <1% in 2015 ranged from 0.15 to 0.87. Assuming testing rates and mass drug administration have remained consistent (twice a year and >80% coverage), linear extrapolation predicted that the three OTZs have some probability of exceeding the elimination threshold by 2030, while two were predicted to have near-zero prevalence. The Tano-Ankobra OTZ, which includes both savanna and forest agro-ecological zones, was a persistent hotspot.

**Conclusions:** This first comprehensive spatio-temporal analysis of onchocerciasis in Ghana reflects the well-documented challenge of onchocerciasis control in forested riverine environments. Persistent transmission foci in eastern and southern Ghana, particularly in forest zones with sustained vector habitat, warrant targeted investigation and intensified intervention to meet WHO elimination thresholds.

## Background

Human onchocerciasis, commonly known as river blindness, is a neglected tropical disease (NTD) that continues to pose public health challenges across sub-Saharan Africa. It is caused by the filarial parasite *Onchocerca volvulus* and transmitted through repeated bites from blackfly vectors (*Simulium* spp.) that breed in fast-flowing rivers and streams (WHO, 2023). The disease derives its common name from both its association with riverine environments and the devastating impact on vision (Biritwum et al., 2021), serving as the second leading cause of infectious blindness after trachoma globally (Gyasi et al., 2023). Onchocerciasis predominantly affects poverty-stricken communities (Chesnais et al., 2017), with 99% of the global cases occurring in 31 endemic countries in sub-Saharan Africa (Coffeng et al., 2013; World Health Organization, 2024). Additional endemic foci exist in Yemen and, to a smaller extent, the Yanomami areas of the Bolivarian Republic of Venezuela and Brazil (WHO, 2025). Beyond the devastating health impacts, onchocerciasis also creates severe socioeconomic burdens. The associated disabilities result in the loss of individuals’ ability to perform daily tasks and work effectively, leading to loss of livelihood and constant dependence on other family members (Otache et al., 2022), perpetuating the cycle of poverty in affected communities.

Efforts to reduce the disease burden began in 1974 with an initial goal of onchocerciasis control through larviciding to reduce blackfly numbers under the operations of the Onchocerciasis Control Programme (OCP), followed by annual and biannual community directed treatment with ivermectin (CDTI) under the African Programme for Onchocerciasis Control (APOC), to reduce the worm burden to a point where blackflies can no longer sustain transmission (Field, Amazigo et al., 2013; Cupp et al., 2011; Hougard et al., 2001; Hougard et al., 1993; Richards et al., 2001; Tekle et al., 2012; Turner et al., 2013). Over the 28 years of the OCP (from 1974 – 2002), 600,000 people were prevented from becoming blind, while a further 18 million children were protected from the risk of infection, and 25 million hectares of arable land were reclaimed. By 2015, with APOC, over 100 million people had been treated, and ∼1 million people did not require treatment (APOC, 2003; CGD, 2004).

Currently, the global estimates in 2017 indicate that approximately 218 million people are at risk of infection and need treatment, with close to 21 million people infected (WHO, 2024). Of these, 14.6 million suffered from skin-associated morbidities and 1.15 million experienced vision impairment in 2017 (WHO, 2022).

Onchocerciasis is endemic in all but one of the 16 administrative regions in Ghana, with only the highly urbanised region of Greater Accra considered non-endemic (Biritwum et al., 2021). Ghana’s extensive river network system provides river basins with suitable environmental conditions ideal for blackfly breeding, as well as local socio-demographic factors that support exposure to blackfly bites. Areas with microfilariae (mf) prevalence requiring CDTI have been categorised into seven operational transmission zones (OTZ) based on river networks: Black Volta, Dayi-Asukwakwa, Oti-Daka, Pra-Offin, Pru-Afram, Tano-Ankobra and the White Volta-Kulpawn (Figure 1B) (WHO, 2016).

**Figure 1.**
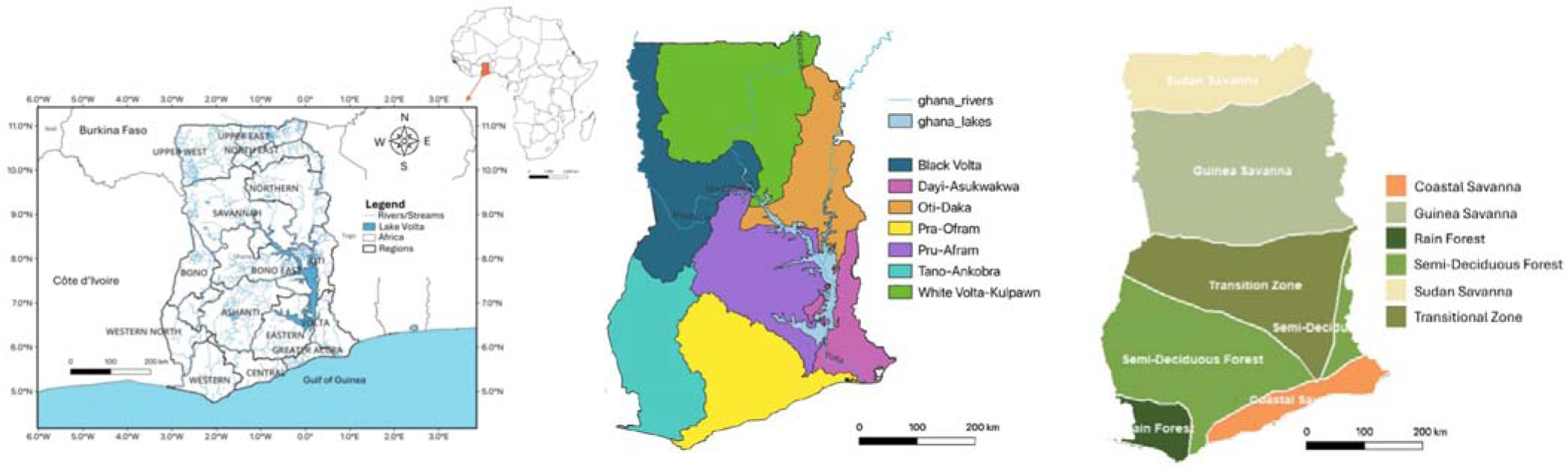
Maps of Ghana. (A) Map of Ghana showing the 16 administrative regions and river network. (B) The seven operational transmission zone (OTZ). (C) Six Agro-ecological zones (Ecozone).

Aerial larviciding was originally focused in the northern regions, with further expansion into more southerly regions in 1987 (World Health Organization, 1987), targeting rivers including the Pra, Pru, Offin, Afram, Bia, and the Bui-Black Volta. The combined efforts of OCP and APOC successfully resulted in a dramatic reduction of mf prevalence in many endemic regions (Biritwum et al., 2021; Frempong et al., 2016). Despite these successes, persistent transmission has been reported in the Pru district of the Bono region along the Pru River basin (Awadzi et al., 2004; Lamberton et al., 2015; Osei-Atweneboana et al., 2007; Kenneth Bentum Otabil et al., 2023). Currently, 5,199,031 people are still considered to be at risk in Ghana (ESPEN, https://espen.afro.who.int/countries/ghana).

The WHO 2030 NTD onchocerciasis roadmap targets elimination, defined as a < 1% skin mf prevalence, in at least one focus in 23 African countries (World Health Organization, 2024). Major challenges to onchocerciasis elimination include civil unrest, cross-border transmission sustained by blackfly and human movement, treatment compliance, drug resistance in the worm population and programme approaches, among others (Ayisi et al., 2024; Babalola et al., 2026; Bush et al., 2018; Gebrezgabiher et al., 2019; Gustavsen et al., 2016; Lakwo et al., 2020; Lakwo et al., 2018; Osei-Atweneboana et al., 2011; Pi-Bansa et al., 2026). More fundamentally, comprehensive information on the spatial distribution of prevalence is required for effective targeted interventions. For instance, the prioritisation for onchocerciasis control was based on identifying high-endemicity special intervention zones (SIZ), where mf prevalence rates exceed 50% (Lamberton et al., 2015; K. B. Otabil et al., 2023), using Rapid Epidemiological Mapping of Onchocerciasis (REMO) (WHO, 2025). Many areas that need interventions to reach elimination, particularly those that are hypoendemic, remain unmapped (Babalola et al., 2026; Isiyaku et al., 2022). Conventionally, surveys are often planned to cover known hyper- and meso-endemic areas, systematically overlooking hypoendemic foci capable of sustaining transmission and historically non-endemic areas that have emerging transmission risks (Babalola et al., 2026; Organization, 2025; Shrestha et al., 2022).

An understanding of the spatial distribution of disease transmission and thus risk of infection will greatly benefit onchocerciasis elimination efforts in Ghana (Babalola et al., 2026), which has a long history of onchocerciasis interventions and features a heterogeneous and ecologically diverse environment (spanning a gradient of savannas and forest agro-ecological zones). Geospatial modelling based on existing epidemiological data, while capturing ecological variation, offers a cost-effective approach to addressing this gap by identifying areas with persistent transmission (persistent hotspots) or potential transmission (emerging hotspots) for resource prioritisation and targeted treatment (O’Hanlon et al., 2016). Spatial models borrow predictive strength from locations with similar climatic and environmental conditions, enabling robust extrapolations to unsampled areas and generating continuous risk maps across wider spatial scales. This allows identification of epidemiologically meaningful spatial units for intervention strategies, while accounting for spatial autocorrelation (Hay et al., 2013; Karagiannis-Voules et al., 2013; Shrestha et al., 2022).

While geospatial modelling has been valuable for transmission delimitation in helminthiasis and vector-borne disease research (Bhatt et al., 2013; Brooker & Clements, 2009; Diggle et al., 2007; Kraemer et al., 2019; Patil et al., 2011; Wang et al., 2024), equivalent tools for onchocerciasis at the national level remain scarce. Spatial models for onchocerciasis have predominantly addressed pre-control periods across continental Africa at low resolution (O’Hanlon et al., 2016; Zouré et al., 2014). These continental-scale approaches may mask crucial local predictors relevant at the country-level transmission dynamics and may not adequately identify the subnational heterogeneity needed to guide interventions. Additionally, some models exclude Ghana entirely (Zouré et al., 2014), and even where prevalence estimates exist for Ghana, the focus is often at a small geographical scale (Barro & Oyana, 2012; Shrestha et al., 2024) or does not include environmental or climatic variables (Biritwum et al., 2021).

Here, we present the first comprehensive spatiotemporal patterns of onchocerciasis mf prevalence distribution across Ghana using 1,353 surveys across 671 villages spanning four decades. We explore the spatio-temporal trends in mf prevalence alongside environmental, bioclimatic and sociodemographic predictors. Our goal is to characterize the spatiotemporal distribution of mf prevalence across Ghana’s major OTZs, identify environmental drivers of persistent transmission hotspots and generate predictive surface maps that can guide mapping surveys and CDTI expansion decisions to support onchocerciasis elimination.

## Methods

### Study area, design and population

Ghana is in West Africa on the Atlantic Ocean, and to the southwest is the Gulf of Guinea. It is bounded by three countries: Burkina Faso, Côte d’Ivoire and Togo to the north, west, and east, respectively, and is divided into 16 administrative regions (Figure 1A). Elevation ranges between 0 and 890 m above sea level (Owusu et al., 2009). As of 2024, the population estimate was 34.43 million (World Bank Group, 2026). Ghana has several freshwater lakes, with the largest man-made lake, Volta, covering 8,502 km^2^ surface area (Figure 1A) (Amisigo et al., 2015; Bessah et al., 2022). Ghana is drained by three major river-basin systems: the Volta Basin, comprising the White Volta, Black Volta, Oti, Lower Volta, and Main Volta; the South-Western Basin, comprising the Pra, Ankobra, Tano, and Bia rivers; and the Coastal Basin, which includes the Kakum, Amissa, Nakwa, Densu, and Ayensu rivers (Agodzo et al., 2023; Amisigo et al., 2015). These form the irrigation sources for the vast agricultural landscape, which covers about 65% of the total land area (Owusu & Waylen, 2012; Wemegah et al., 2020), and generates ∼30% of the country’s GDP (Amisigo et al., 2015). The Ghana Health Service has further delineated the country into seven operational onchocerciasis transmission zones (OTZs) based largely on river-basin systems and associated transmission patterns (Figure 1B). The Black Volta, White Volta–Kulpawn, and Oti–Daka OTZs are located primarily in northern Ghana; the Dayi–Asukawkaw OTZ is situated in the east; the Pru–Afram OTZ spans the central part of the country; the Pra–Offin OTZ extends from central Ghana towards the southern coast; and the Tano–Ankobra OTZ encompasses areas in western and south-western Ghana. Together, these zones provide an operational framework for describing the geographical distribution of onchocerciasis transmission across Ghana and serve as the principal units for planning and implementing control, surveillance, and elimination activities (Biritwum et al., 2021). Ghana is divided into six agro-ecological zones (Figure 1C): Sudan and Guinea savannas in the north, a “Transitional” zone in the middle, semi-deciduous forest in the south and east, rainforests in the southwest, and coastal savanna along the southern ocean border (Darko et al., 2023). The climate is tropical, with two main seasons: rainy and dry.

### Data Sources

#### Onchocerciasis infection data

Onchocerciasis mf prevalence data, with geocoordinates spanning 1975 to 2015, were obtained from the NTDs Programme of the Ghana Health Service and the publicly available Expanded Special Project for Elimination of Neglected Tropical Diseases (ESPEN) database portal. The data were collected as part of the REMO to identify high-risk areas for onchocerciasis control implementation (Noma et al., 2013). Both data sets were combined and manually checked for duplicates to obtain a consensus data file. Discrepancies between geocoordinates were also verified via Google Maps (Google Inc., 2026), and observations with unresolved coordinates were removed from the dataset.

The data used for analysis were the community prevalence; i.e., the proportion of community members testing positive, conditional on the total number tested at village clinics. Thus, while the geographical location of the testing clinic was known, we lacked individual-level data on participants’ home location, which is the proximal determinant of infection risk. To estimate the relative exposure to causal factors for participants tested at each village clinic, we used a hypothetical ‘catchment area’. This was defined as a 10x10 km square sampling window centred on the location of the clinic, which was used to quantify covariates thought to influence infection rates (see ‘Modelled predictors of variation in infection rates’ below). These sampling dimensions were based on the ‘5 km from watercourses’ linear threshold distance used to identify first-line villages, which are considered to have high infection risk (Boakye et al., 2023; Cromwell et al., 2021). We assumed participants might travel within 5 km to the clinic from any direction, which was considered a robust likely maximum travel distance based on field experience in rural Ghana, but would not be accurate if any participants travelled to the clinic from a longer distance for testing.

#### Climatic and Environmental data

Climatic and explanatory environmental data that are expected to influence mf prevalence were identified based on the literature and were obtained through the Google Earth Engine platform (Gorelick et al., 2017). These include climatic variables (rainfall and temperature), bioregional land use (landcover types, urbanisation, and agro-ecological zones), environmental characteristics (soil moisture, elevation, flow accumulation, direction of water flow, distance to nearest river and vegetative index), and socio-demographic data (population density) (Supp Table 1).

For all predictors, we sampled conditions within each of the 10 x 10 km village sampling window, selecting a time closest to the mid-year of survey data duration if the sample collection month was not recorded. For continuous predictors (raster layers), we used the mean cell value within the window; for categorical predictors, the relevant factor level (Table 1) within the window was used (note, no categorical variables had more than a single factor level within each village sampling window). Prior to all analyses, geocoordinates were reprojected to WGS 84 / UTM zone 30 N, EPSG:32630 (easting and northing) for accurate measurements of distances and to prevent computational errors.

**Table 1.** Factor levels for spatial units used in analysis of correlations between epidemiological and ecological variables associated with onchocerciasis prevalence and distribution in Ghana.

| **Spatial Unit** | **Level** |  |  |  |  |  |  |  |
| --- | --- | --- | --- | --- | --- | --- | --- | --- |
| **Operational Transmission Zone** | Black Volta | White Volta-Kulpawn | Oti-Daka | Pru-Afram | Dayi-Asukawkaw | Pra-Offin | Tano-Ankobra |  |
| **Ecozone** | Sudan Savanna | Guinea Savanna | Transitional Zone | Semi-Deciduous Forest | Rain Forest | Coastal Savanna | Rain Forest and Semi-deciduous forest |  |
| **Urbanization** | Very Low Density Rural | Low Density Rural | Rural Cluster | Suburban or Peri-urban | Semi-dense Urban Cluster | Dense Urban Cluster | Urban Cluster | Water |
| **Landcover** | Croplands | Grasslands | Savannas | Woody Savannas | Forest-Built | Mosaic croplands/natural vegetation | Water |  |

### Data analysis

Our overall goal was to understand spatio-temporal variation in onchocerciasis prevalence across Ghana using two different approaches: (1) summarising empirical patterns for different administrative boundaries and different periods of time, and (2) modelling prevalence rates within a hierarchical Bayesian generalised additive model (GAM) framework while accounting for ecozone-specific intervention responses and residual spatial heterogeneity as a function of covariates that we anticipated would be influential (climate, hydrology, population density, and bioregional land use).

#### Point-estimates of the spatio-temporal distribution of onchocerciasis microfilariae prevalence

To illustrate variation in spatial data collection and prevalence rates based purely on empirical patterns in the data, we first present prevalence organised by different administrative or ecologically defined reporting units. This indicates how collated prevalence information is conditional on the spatial reporting units. The mf prevalence rates were estimated for spatial units defined by administrative districts, transmission zones, agro-ecological zones, landcover, and urbanisation, calculated as the total number of mf-positive individuals detected in the geospatial unit of interest, divided by the total number examined across all spatial units.

#### Modelled predictors of variation in prevalence rates

To model the effect of four hypothesised major influences on prevalence rates, we used a combination of individual spatial layers and composite predictors integrating multiple source datasets. The latter were used to reduce collinearity among individual covariates and improve interpretability within a spatial modelling framework while still capturing variability in the strength of distinct ecological and socio-environmental processes underlying transmission. These four categories were:

1. **Climatic suitability.** Hypothesised effect: Climate is expected to be positively correlated with prevalence. Vector development and activities are heavily dependent on temperature. Temperature also regulates parasite development in vectors as well as in humans. Hence, optimal average temperatures and low temperature variability are expected to enhance transmission efficiency and, consequently, increase prevalence (Takaoka et al., 1982).
2. **Probability of flowing water.** Hypothesised effect: Blackfly vectors breed in fast-flowing rivers. As such, areas combining moderate slopes, sufficient rainfall, and high surface wetness are hypothesised to support sustained transmission (Shrestha et al., 2024; Shrestha et al., 2022).
3. **Probability of flies obtaining a blood meal.** Hypothesised effect: unimodal (hump-shaped) response with population density. At low population densities, increasing host availability enhances vector–host contact, increasing prevalence. However, reduced vector habitat and improved infrastructure are often associated with high population densities (e.g. major urban centres), prevalence is expected to decline, reflecting a balance between host availability and environmental suitability for transmission (Basáñez, 1994; Basáñez et al., 1995; Cheke et al., 2017).
4. **Bioregional, vegetation, and land-use context.** Hypothesis: Bioregional and land-use characteristics influences microclimate, vector habitat suitability, and human-to-environment interactions. Natural and semi-natural landscapes are hypothesised to support a higher prevalence relative to heavily modified or urbanised land-use types. This predictor also coincides with differences in the distribution of different species of black flies (Ayisi et al., 2022; O’Hanlon et al., 2016), which could also impact prevalence rates. Agro-ecological zones were assigned using published data (Darko et al., 2023; Mensah et al., 2026).

Composite predictors for each of the four major influences were developed from continuous spatial layers in two phases. First, we standardized all predictors and then used a linear forward selection approach in R v4.5.2 (Posit team, 2025; R Code Team, 2025) as implemented in *adespatial::forwardsel v0.3.28* (Blanchet et al., 2008) with α set to 0.2. This reduced the number of candidate environmental covariates, which we then combined using principal components analysis (PCA) and extracted the first principal component to capture the major axis of variation associated with all predictors within the composite to represent their influence in the model.

### Modelling approach

All models were fit using the *brms* package *v2.23.0* (Bürkner, 2017) in R *v4.5.2* and estimated via Hamiltonian Monte Carlo using the No-U-Turn Sampler implemented in Stan (Carpenter et al., 2017). Weakly informative regularizing priors were specified for all model parameters. Four independent Markov chains were run for 4,000 iterations each, with the first 2,000 iterations discarded as warm-up and sampling parameter adapt delta (determining the step size in the sampler) to 0.99, 15 maximum tree depth and a total of 8,000 post-warmup draws. Weakly informative prior distributions were specified for all model parameters and smoothness parameters were estimated using the Bayesian penalisation framework implemented in *brms* (Bürkner, 2017) (see ‘Model convergence and diagnostics’ below).

#### Model

The response outcome modelled was the observed count of mf-positive individuals (Positive) from a binomial denominator of individuals examined (Examined) at each survey site, with the linear predictor specified on the logit scale. Because the data contained a high proportion of zero-prevalence observations, particularly in later records, we used a zero-inflated beta-binomial model (Hu et al., 2018; Wen et al., 2024). While a hurdle model would offer an alternative approach to dealing with excess zeros (Lambert, 1992; Mullahy, 1986). However, individual ivermectin use histories were unavailable, limiting our ability to distinguish structural zeros attributable to treatment from zeros arising from other causes. We also preferred a binomial response distribution as it allowed us to include information on the number of tests undertaken in each village rather than model the proportion of positive cases. Ultimately, a zero-inflated beta-binomial distribution was applied to account for both excess zeros and overdispersion, proving superior relative to a standard binomial model (beta-binomial model weight > 0.99). Zero inflation accommodates surveys in which no infections were observed beyond what would be expected under binomial sampling alone. The beta-binomial component further allows prevalence to vary among surveys due to unobserved heterogeneity in infection risk, providing a flexible and biologically realistic representation of parasitological survey data.

Variation in mean prevalence and zero inflation were modelled using penalised thin plate regression splines (smoothing functions) implemented in a Bayesian framework (Bürkner, 2017; Miller, 2025; Wood, 2003, 2025). The zero-inflation component was modelled as a function of a single covariate, time (year). Mean prevalence rates were explored using several candidate models with different combinations of linear and non-linear predictors, the latter implemented using both global and factor smooths of the predictors, following the framework for generalised additive models in Pedersen and colleagues (Pedersen et al., 2019) before adopting a final model. Model selection proceeded by comparing different combinations of predictors across six candidate models. “*Leave-one-out*” cross-validation (LOO-CV) was applied to assess the predictive performance and to compare candidate models (see Table S5), with the final model selected based on comparing stacked model weights and theoretical parsimony (Vehtari et al., 2017).

The final population level (aka fixed) effects mean response model structure included five linear and four non-linear terms. Linear terms included two continuous (wetness and population density) and three factors (ecozone, urbanization, and landcover). Non-linear terms were global smoothers for year and probability of runoff (ro_pca), a factor smooth of year (estimating variation from the global smoother for each level of a factor variable) for OTZ and a two-dimensional tensor product smoother of X-Y coordinates to account for spatial autocorrelation. A group level (aka random) effect was included for village to account for temporal autocorrelation. However, because sampling was not consistent within and among villages, village-level effects were nested within ecozones to improve the ability of the model to provide robust estimates of spatial trends. The complete model structure follows.

The mean response (*µ_ijkt_*) was modelled as:

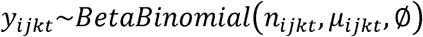

Where:

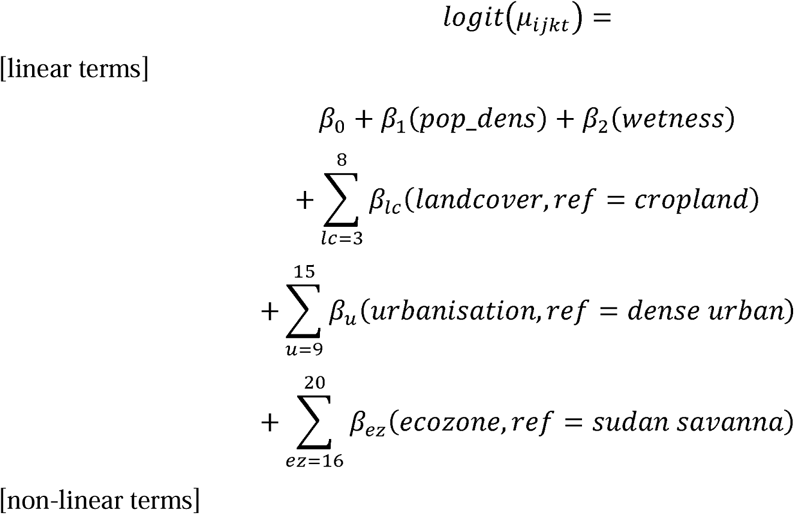

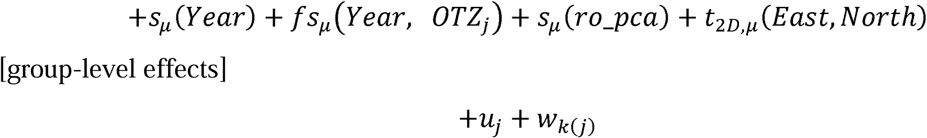

The zero-inflation component (*π_ijkt_*) was modelled as:

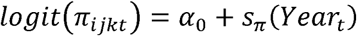

Prior distribution specifications for model parameters were:

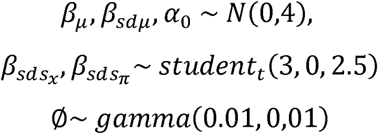

Where:

*y_ijkt_* is the number of positive tests *i*, in village *k*, ecozone *j*, and year *t*.

*n_ijkt_* is the number of trials in village *k* in year *t*.

*µ*_ijkt_ is the mean expected proportion of positive tests.

*β*_0,_ *α*_0_ global intercepts for mean and zero-inflated response.

*s_µ_*(*pred*) is a smoothing function for continuous predictor ‘*pred*’, to estimate the mean response, each with two parameters a linear spline component and a deviation (wiggliness) component (notated *β_sµ_* and 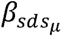 respectively). The equivalent smoothing function for zero-inflation is denoted with subscript π.

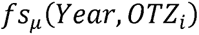 is a factor smoothing function, estimating deviations in the mean prevalence from the global smoother for each operational transmission zone.

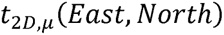 is a two-dimensional tensor product smoothing term of the Easting and Northing coordinates for each village. This is included to control for spatial autocorrelation in prevalence rates.

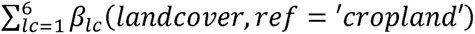 coefficients for level of categorical variables (here landcover).

*u_j_*, *w_k_*_(*j*)_ are random intercepts for village *k*, nested in ecozone *j*.

∅ is the precision parameter controlling variance of the non-zero component.

#### Model convergence and validation

Model convergence was assessed using Gelman-Rubin diagnostic (R-hat) and effective sample sizes (ESS). Visual inspection of trace plots generated using the *“mcmc_trace”* function in the *brms* and *bayesplot* (Gabry & Mahr, 2025) packages in R showed good mixing and reliable sampling. Model adequacy was evaluated using posterior predictive checks and Bayesian R^2^ (Gelman et al., 2013; Gelman et al., 2019). For comparison purposes, observed data with Wilson confidence intervals were overlaid on the fitted values and plotted using the R package *ggplot2 v4.0.1* (Wickham, 2016).

#### Posterior predictive prevalence distributions and future projection

Posterior predictive distributions (PPD) for the period 1975–2015 were generated for each OTZ from the final model. For OTZ that overlapped more than one ecozone, we made individual predictions for each of the latter. For these predictions, all continuous covariates were fixed at their mean values, while the number examined was held constant at 100. Predictions were generated using a seed value of 108 and based on 1,000 posterior draws from the fitted model.

Two types of PPD were generated. First, we created a smooth time series over the modelled period (and beyond – see below) for each OTZ to provide an indication of variation in temporal dynamics. Second, spatial prediction surfaces were subsequently generated across Ghana using a regular prediction grid. Grid cells were spaced at approximately 1.6 km in the east–west direction and 2.4 km in the north–south direction, corresponding to a nominal spatial resolution of approximately 2 km. Predictions at each grid cell were obtained using the fitted Bayesian zero-inflated beta-binomial geostatistical model and subsequently clipped to Ghana’s national district boundaries for visualization. Prediction uncertainty was quantified using the posterior 95% credible intervals, represented by the lower (2.5th percentile) and upper (97.5th percentile) bounds of the posterior predictive distributions.

Although we acknowledge the uncertainties in any extrapolation beyond the temporal range of the fitted data (particularly for models that include smoothing terms), prevalence trends were extended to 2030 to provide a possible future trajectory. This was considered especially important given the period of time that has already elapsed since the last survey data available. Owing to our use of thin plate regression splines, these extrapolations essentially amount to linear projections from the end of the data series. Assuming that all covariates and current intervention effort and spatial prioritisation remain constant, we believe this provides a reasonable basis to project prevalence. However, we stress these projections should be interpreted as indicative conditional scenarios rather than definitive forecasts.

One of our aims for the estimated PPD was to assess the probability that prevalence rates had reached the World Health Organization elimination threshold of 1% mf prevalence. For both 2015 and 2030, we calculated the median prevalence as an indication of centrality in the predicted distribution and quantified the probability that the 1% threshold had been achieved based on the amount of probability density in the PPD that was below this value using the ‘hypothesis’ function in *brms*. That is, a PPD with 0.8 below 0.01 was interpreted as 80% confidence that the 1% threshold had been reached. This posterior probability of elimination was estimated for OTZ, ecozone, national, and district grids. The spatial units were classified using the same approach, according to their posterior support for elimination, with high posterior probabilities indicating strong evidence of achieving the elimination target and low posterior probabilities indicating evidence of likely failure to reach the threshold. Conversely, the probability of failing to achieve elimination was calculated as the proportion of posterior draws with predicted prevalence equal to or exceeding 1%.

Decision thresholds were interpreted probabilistically, such that posterior probabilities exceeding 0.95 were considered strong evidence of elimination, probabilities below 0.05 indicated strong evidence of failure to achieve elimination, and intermediate probabilities reflected substantial uncertainty regarding elimination status. Thus, posterior probabilities provide a formal Bayesian framework for quantifying uncertainty around progress toward elimination and identifying areas where intensified intervention may be required.

National and district-level predictive spatial maps were generated with all continuous predictors (ro_pca, TCW_wetness, ln_pop) held at mean values, while the categorical spatial units were set to their reference values (OTZ = Black Volta, Ecozone = CS, Urbanization = LDR and Landcover = Cropland/Natural Vegetation Mosaics).

## Results

### Data summary

#### Spatial distribution of onchocerciasis microfilarial prevalence

A total of 1,353 onchocerciasis mf prevalence data points from 671 villages in 147 districts were included in the analysis. A total of 221,521 individuals were sampled from 1975 to 2015, and out of these, 45,709 tested positive, for an overall prevalence across Ghana of 20.6% (95% CI: 20.5-20.8). At the village level, the number of individuals sampled per village ranged from 10 to 1134, with prevalence ranging from 0 to 88.9%. Prevalence across the country was heterogeneous, and many districts did not have any villages assessed for mf prevalence at any time (Figure 2A&B). Survey efforts changed over time, resulting in patchy data across much of the country at any given time point (Figure 2).

**Figure 2.**
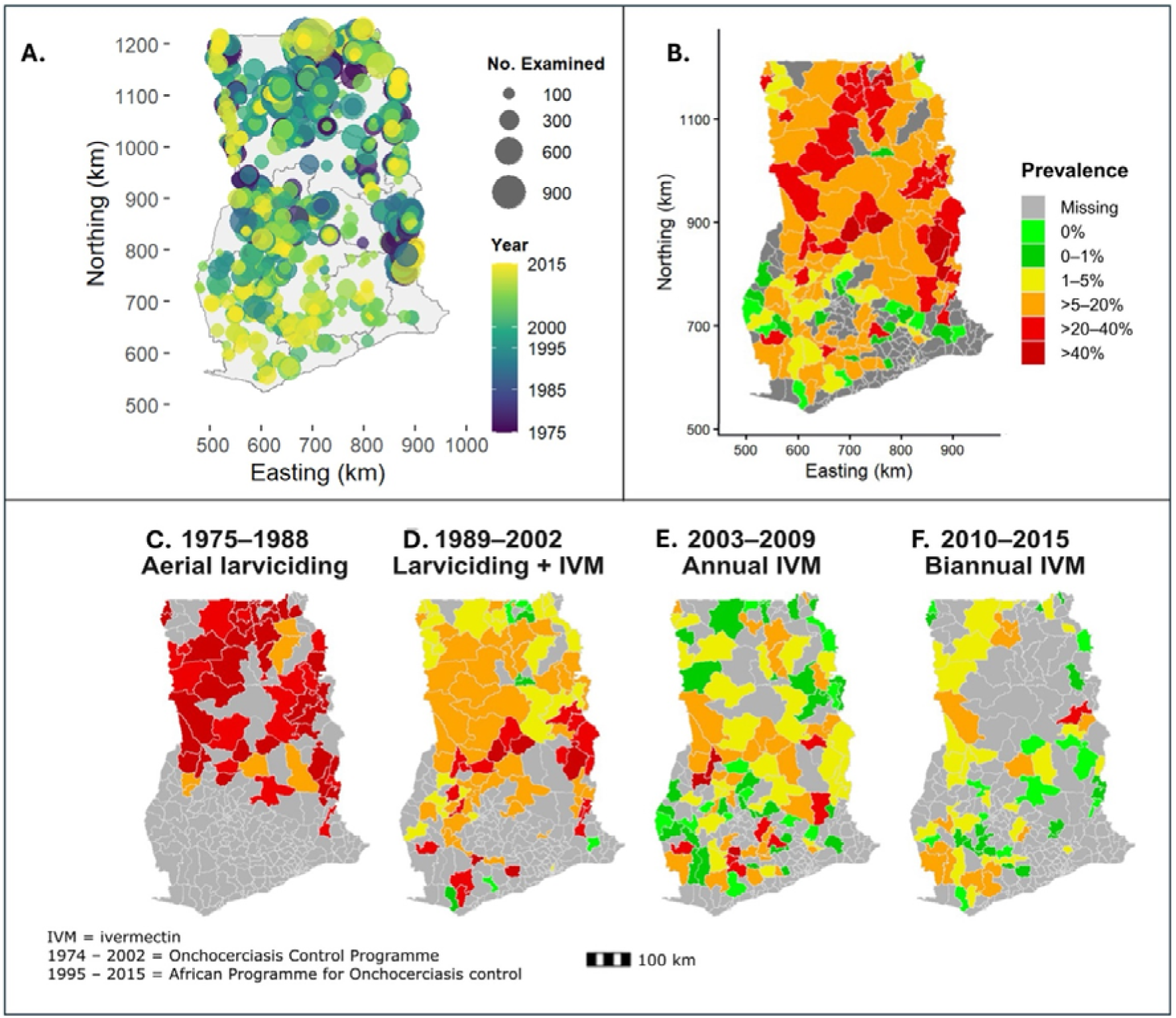
Spatial distribution of raw onchocerciasis microfilariae (mf) data in Ghana 1975 to 2015. Panel A shows the location of all sampling sites, with point size proportional to the number of individuals examined and colour indicating the survey year, with dark blue representing the earliest survey years and yellow denoting the latest years. Panel B shows the pooled mf prevalence (total positive / total examined) aggregated by districts across all the years. C to D, are the district-level mf prevalence under four intervention phases. (C) From 1974 to 1988, when, aerial larviciding to control blackfly numbers was the main control strategy, (D) From 1989 to 2002, when both aerial larviciding and the early introduction of community-directed treatment with ivermectin (CDTi) were implemented. (E) 2003 – 2009, marked by the large-scale rollout of annual CDTi. (F) 2010 – 2015, when biannual CDTi was introduced in certain communities with high transmission. Grey areas represent districts with no survey data.

Prevalence across districts was heterogeneous over time (Figure 2C-F). The peak prevalence of 73.8% was recorded in 1988, but this value is likely biased because>90% of the tests (1,571) came from the hyperendemic Pru-Afram OTZ. The highest reliable prevalence (∼65%) was recorded between 1975-1976 (see Figure 2C). There was a gradual decline throughout the 1980s when aerial larviciding was underway, then a sharp drop in prevalence with the introduction of ivermectin in 1993 (Figure 2D). Prevalence stabilised at low levels (∼5 to 10%) under annual CDTI during APOC (Figure 2E). When biannual CDTI was implemented in certain areas in 2009, prevalence dropped further to very low levels, almost approaching zero (0.61%) (Figure 2F).

### Model Convergence

All model parameters showed excellent convergence (RC = 1.00), with bulk and tail effective sample sizes of 2,463 and 5,448 well above the recommended minimum (Vehtari et al., 2021) showing excellent sampling efficiency and zero divergent transitions across 8,000 post-warmup samples. The overdispersion parameter (φ = 16.14, 95% CrI: 13.62 – 18.89) confirmed substantial extra-binomial variation consistent with clustered transmission dynamics and heterogeneous survey conditions across sites and years. The zero-inflation probability was negligible (zi = 0.01, 95% CrI: 0.00 – 0.02), indicating that structural zeros were rare and that near-zero prevalence observations in the post-intervention period were adequately accommodated by the beta-binomial likelihood with the zero-inflation component. The model predictions were strongly correlated with observed prevalence (Pearson’s *r = 0.947*, *R^2^=0.897*and *p = 0.01)*, with root mean square error (RMSE) of 0.072 and average absolute prediction error (MAE) of 0.049 (Supp Figure 2), indicating only a 7.2% points difference between the observed and fitted prevalence on average. The dense cluster of points near the origin reflects the high proportion of near-zero prevalence observations in the post-intervention period. In addition, the Pareto-smoothed importance sampling leave-one-out diagnostics indicate that most of the observations were well behaved (94.1% within *k* ≤ *0.7.* However, 80 observations (5.9%) had *k* ≥ *0.7*, while four observations (0.3%) had *k > 1* (Table S4), suggesting a small number of influential observations.

### Modelled onchocerciasis mf predicted prevalence in Ghana

Of the four categories of influencing variables, the composite predictors and first principal component of the PCA were: (1) Climate composite predictor combining mean temperature and isothermality (bio3), quantified as the first principal component including temp_avg and temperature variability; (2) Hydrology composite predictor combining Topography (slope + elevation) + Climate (precipitation of wettest quarter (bio16)) + (temperature / rainfall covariance (bio8)) + Soil moisture and hydrology (TCW_wetness + Flow_accummulation), with first principal component including: slope, elevation, precipitation of wettest quarter (bio16), temperature/rainfall covariance (bio8), Tasselled cap wetness, and flow accumulation; (3) Blood meal availability, or population density in Ghana from 1975-2015, quantified as the log-transformed mean population density within each 10 x 10 km buffer around each point from the 2000 census; (4) for Bioregional land use, landcover variable and urbanisation were used, while the highly correlated vegetative indices (Figure.S1): Normalized Difference Vegetation Index (NDVI) and Enhanced Vegetation Index (EVI) were excluded. Finally, the climate composite predictor was excluded because it was highly correlated with geographical location (easting and northing) (See Supp Figure 1).

Predicted mean posterior prevalence draws show a decline from around 53.3 (95% CrI: 31.9% – 57.8%) to 39.3% (295% CrI: 8.6% - 51.5%) from the mid-70s to the mid-80s during the OCP era, when aerial larviciding was the main control strategy. The license of IVM and subsequent mass rollout led to a dramatic decline in prevalence. Prevalence levels fall below 5% by 2015, reflecting the accelerated impact of ivermectin scale-up under the APOC/CDTI from the early 2000s (Figure 3). The wide credible intervals (grey shading) in the early survey years before 1985 indicate the relatively sparse data, which narrow as more data become available (Figure 3).

**Figure 3.**
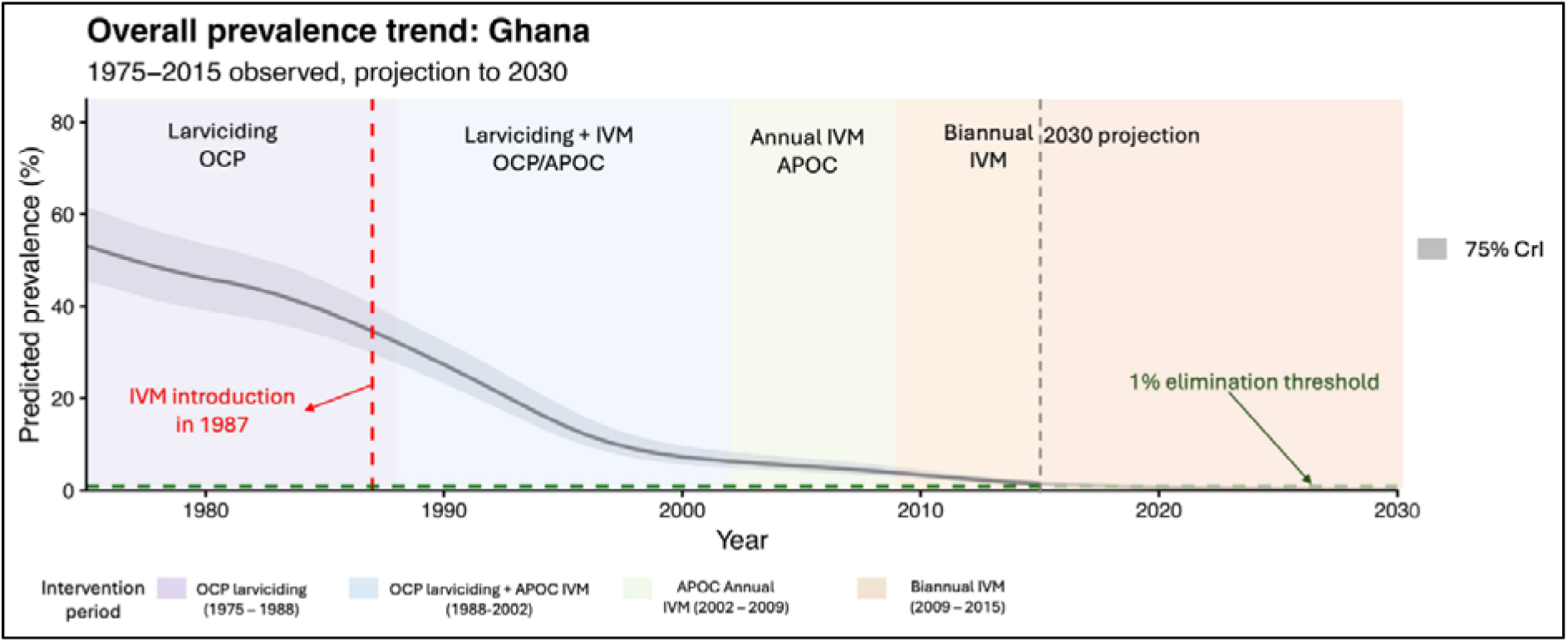
Modelled temporal trend in predicted prevalence in Ghana across survey years and projections to 2030. The grey sigmoid shows prevalence decline, and the red dashed line indicates the point when ivermectin was licensed for human use and piloted in hyperendemic areas. The WHO 1% elimination threshold is indicated in a green horizontal dashed line. The grey vertical dashed line is the 2015 mark, beyond which linear projections are made assuming testing rates and community directed treatment with ivermectin remain consistent with the modelled period since 2015.

Among the environmental predictors (Supp Figure 4), prevalence increased modestly with increasing runoff probability (ro_pca) but plateaus (Supp Figure 4A), suggesting that, beyond a threshold of hydrological suitability, additional runoff does not increase transmission any further. Hydrological conditions associated with sustained river flow may enhance vector breeding suitability. Surface wetness (TCW_wetness) showed a strong positive relationship with predicted prevalence (Supp Figure 4B, where wetter landscapes are associated with increased transmission, consistent with *Simulium* breeding dependence on moist riverine environments. Population density showed no independent effect on predicted prevalence, suggesting limited explanatory contribution once environmental and spatial factors (such as urbanisation and landcover category) were accounted for. Differences in predicted prevalence across urbanisation classes were modest, with substantial overlap in posterior credible intervals, indicating limited evidence for strong independent effects. Very low density rural (VLDR) settings, however, showed the highest predicted prevalence, followed by rural clusters (RU). The OTZs showed marked variation, consistent with persistent spatial heterogeneity in transmission intensity after adjusting for temporal effects, with the Dayi-Asukawkaw OTZ showing the lowest estimates, while the Pra-Offin and Tano-Ankobra OTZs had high prevalence. This is further supported by the posterior estimates, which indicated positive associations between prevalence and two types of land cover (woody savannas and savanna), TCW, and VLDR settlements. Several contrasts between ecozones and urbanisation had credible intervals that overlapped zero, indicating substantial uncertainty in their effects after accounting for other covariates (Supp Figure 5).

### Spatio-temporal trends in predicted prevalence across spatial units

Spatially explicit posterior probabilities of achieving the 1% elimination threshold by 2030 are presented in Figure 4, highlighting heterogeneous progress toward elimination across Ghana. By 2015, the median prevalence in OTZ in Ghana was estimated to be between 0.43 and 1.97 (Table 2). The probability that prevalence rates at the OTZ level were less than 1% in 2015 ranged from 0.15 to 0.87. Assuming testing rates and CDTI have remained consistent with the modelled period since 2015, linear extrapolation predicted that the median prevalence will range from 0 – 0.26, with a greater than 0.87 probability that prevalence rates throughout Ghana would be below 1% by 2030 (Figure 4). The White Volta-Kulpawn, Black Volta, Dayi-Asukawkaw and Tano-Ankobra OTZs, however, are most likely to retain some probability of exceeding 1%, while Pra-Offin and Pru-Afram are predicted to have near-zero prevalence. Similarly, median values were seen in Ecozones in 2015 (0.45 and 1.97), with probabilities slightly lower, 0.13 to 0.86 (Supp Table 5). In 2030, the median prevalence was between 0 and 0.24, with northern Savannas (within the White Volta-Kulpawn OTZ) and Rain Forest ecozones (in the Tano-Ankobra OTZ) having the lowest probability of prevalence rates being less than 1% (Supp Table 5).

**Figure 4.**
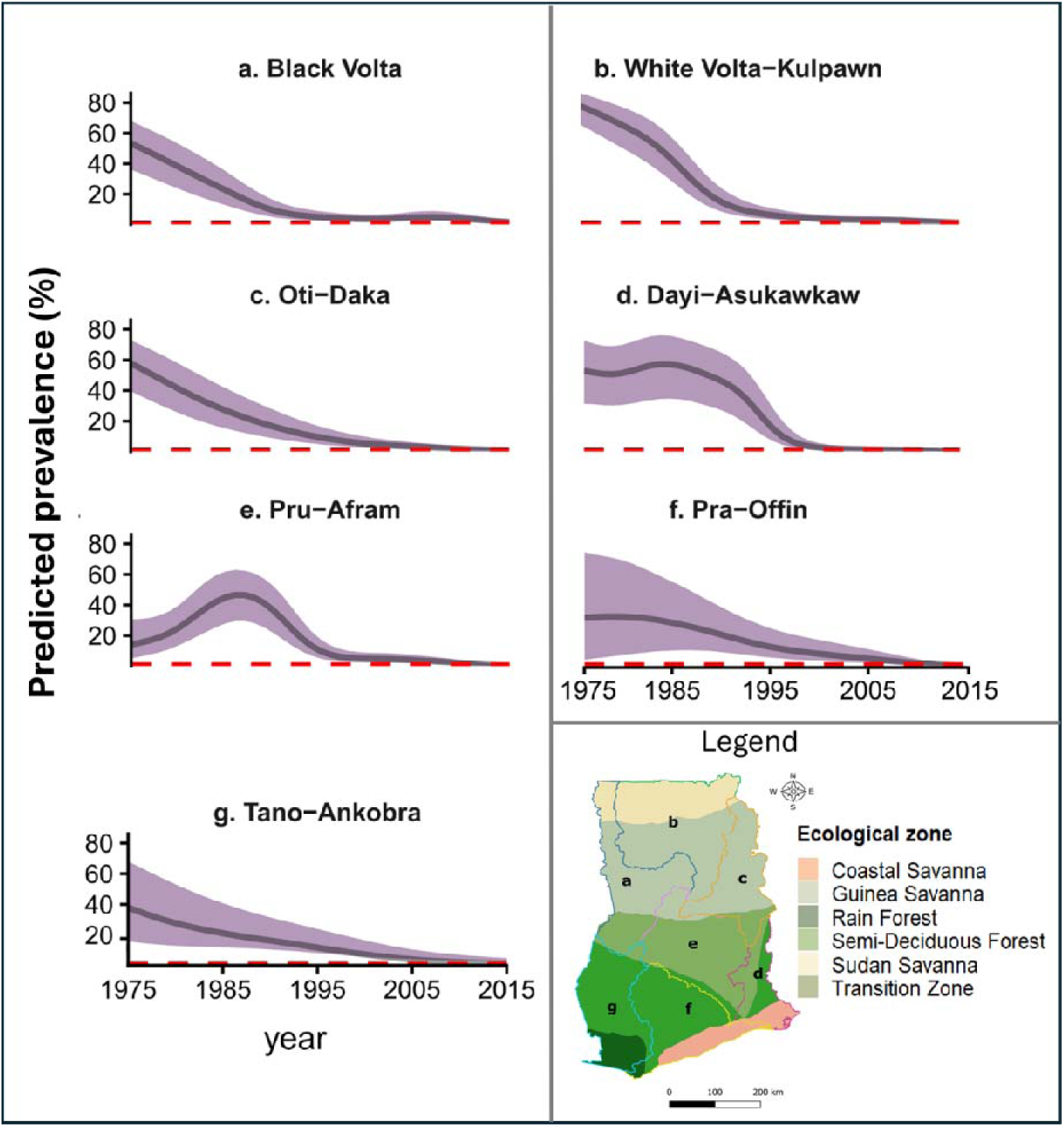
Modelled temporal trends in prevalence for OTZ in Ghana from 1975-2015. OTZ vary in the representation of different ecozones, which is depicted in the Legend (letters correspond to OTZ in individual panels, shading shows ecozone boundaries. Map of agro-ecological zones in Ghana generated in QGIS 3.40.0 using river boundaries described by Darko and colleagues (Darko et al., 2023).

**Table 2.**
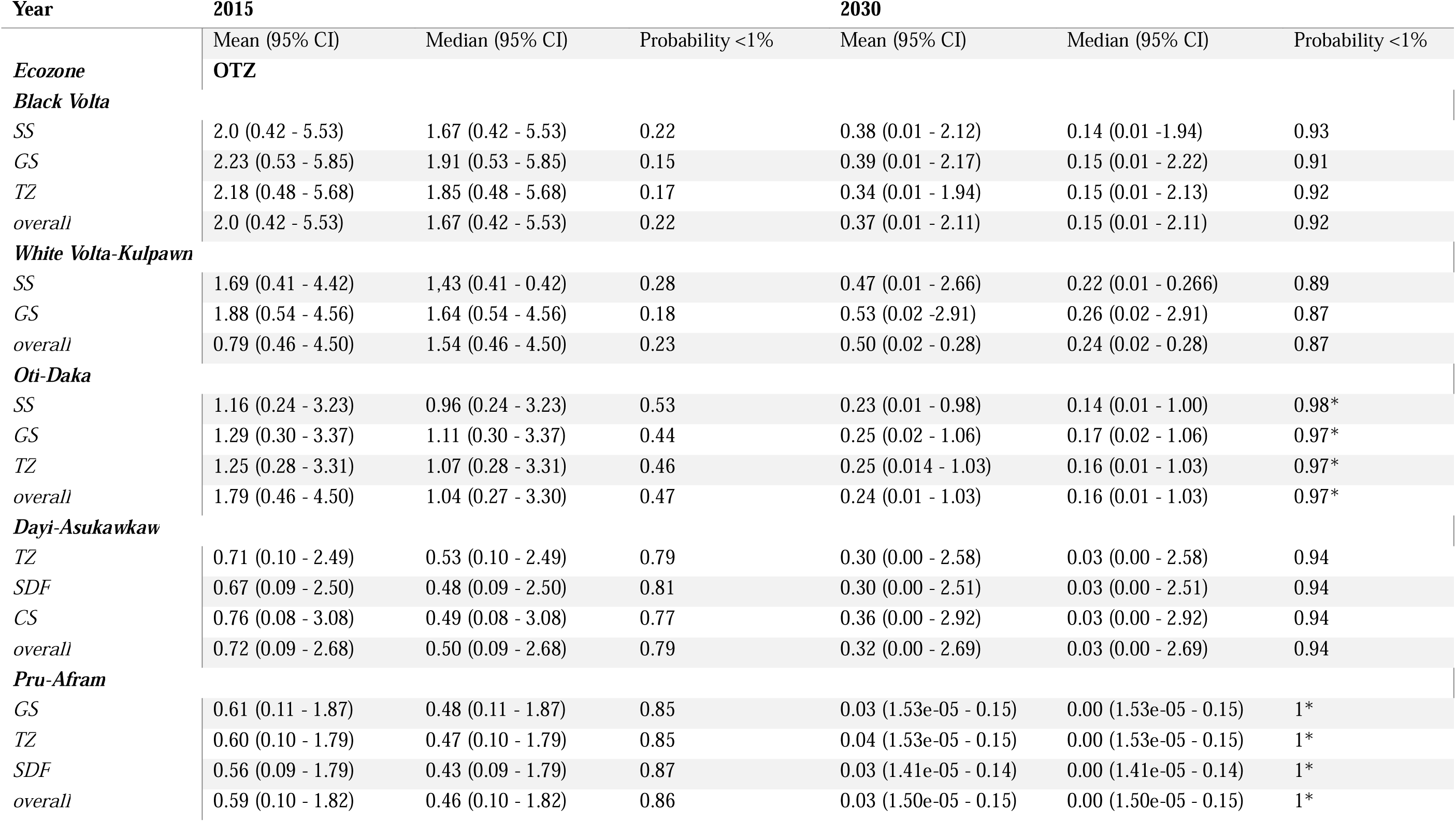
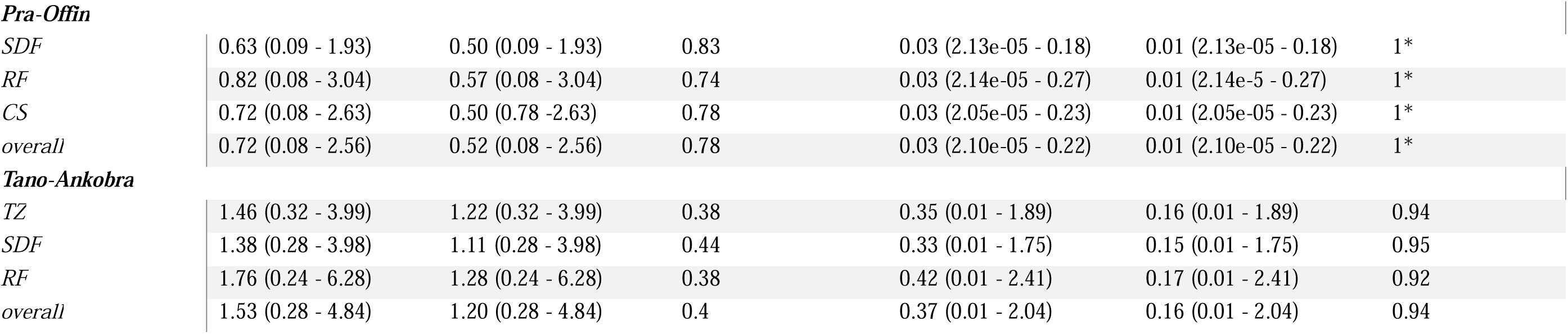
Posterior probabilities of achieving the World Health Organization 1% onchocerciasis elimination threshold by 2015 and 2030 for ecozones within Operational Transmission Zones (OTZ). SS: Sudan Savanna; GS: Guinea Savanna; TZ: Transitional Zone; SDF: Semi-Deciduous Forest, CS: Coastal Savanna; RF: Rain Forest. * probability >95% of achieving elimination.

| Year | 2015 |  |  | 2030 |  |  |
| --- | --- | --- | --- | --- | --- | --- |
|  | Mean (95% CI) | Median (95% CI) | Probability <1% | Mean (95% CI) | Median (95% CI) | Probability <1% |
| **Ecozone** | **OTZ** |  |  |  |  |  |
| **Black Volta** |  |  |  |  |  |  |
| SS | 2.0 (0.42 - 5.53) | 1.67 (0.42 - 5.53) | 0.22 | 0.38 (0.01 - 2.12) | 0.14 (0.01 - 1.94) | 0.93 |
| GS | 2.23 (0.53 - 5.85) | 1.91 (0.53 - 5.85) | 0.15 | 0.39 (0.01 - 2.17) | 0.15 (0.01 - 2.22) | 0.91 |
| TZ | 2.18 (0.48 - 5.68) | 1.85 (0.48 - 5.68) | 0.17 | 0.34 (0.01 - 1.94) | 0.15 (0.01 - 2.13) | 0.92 |
| overall | 2.0 (0.42 - 5.53) | 1.67 (0.42 - 5.53) | 0.22 | 0.37 (0.01 - 2.11) | 0.15 (0.01 - 2.11) | 0.92 |
| **White Volta-Kulpawn** |  |  |  |  |  |  |
| SS | 1.69 (0.41 - 4.42) | 1.43 (0.41 - 0.42) | 0.28 | 0.47 (0.01 - 2.66) | 0.22 (0.01 - 0.266) | 0.89 |
| GS | 1.88 (0.54 - 4.56) | 1.64 (0.54 - 4.56) | 0.18 | 0.53 (0.02 - 2.91) | 0.26 (0.02 - 2.91) | 0.87 |
| overall | 0.79 (0.46 - 4.50) | 1.54 (0.46 - 4.50) | 0.23 | 0.50 (0.02 - 0.28) | 0.24 (0.02 - 0.28) | 0.87 |
| **Oti-Daka** |  |  |  |  |  |  |
| SS | 1.16 (0.24 - 3.23) | 0.96 (0.24 - 3.23) | 0.53 | 0.23 (0.01 - 0.98) | 0.14 (0.01 - 1.00) | 0.98* |
| GS | 1.29 (0.30 - 3.37) | 1.11 (0.30 - 3.37) | 0.44 | 0.25 (0.02 - 1.06) | 0.17 (0.02 - 1.06) | 0.97* |
| TZ | 1.25 (0.28 - 3.31) | 1.07 (0.28 - 3.31) | 0.46 | 0.25 (0.014 - 1.03) | 0.16 (0.01 - 1.03) | 0.97* |
| overall | 1.79 (0.46 - 4.50) | 1.04 (0.27 - 3.30) | 0.47 | 0.24 (0.01 - 1.03) | 0.16 (0.01 - 1.03) | 0.97* |
| **Dayi-Asukawkaw** |  |  |  |  |  |  |
| TZ | 0.71 (0.10 - 2.49) | 0.53 (0.10 - 2.49) | 0.79 | 0.30 (0.00 - 2.58) | 0.03 (0.00 - 2.58) | 0.94 |
| SDF | 0.67 (0.09 - 2.50) | 0.48 (0.09 - 2.50) | 0.81 | 0.30 (0.00 - 2.51) | 0.03 (0.00 - 2.51) | 0.94 |
| CS | 0.76 (0.08 - 3.08) | 0.49 (0.08 - 3.08) | 0.77 | 0.36 (0.00 - 2.92) | 0.03 (0.00 - 2.92) | 0.94 |
| overall | 0.72 (0.09 - 2.68) | 0.50 (0.09 - 2.68) | 0.79 | 0.32 (0.00 - 2.69) | 0.03 (0.00 - 2.69) | 0.94 |
| **Pru-Afram** |  |  |  |  |  |  |
| GS | 0.61 (0.11 - 1.87) | 0.48 (0.11 - 1.87) | 0.85 | 0.03 (1.53e-05 - 0.15) | 0.00 (1.53e-05 - 0.15) | 1* |
| TZ | 0.60 (0.10 - 1.79) | 0.47 (0.10 - 1.79) | 0.85 | 0.04 (1.53e-05 - 0.15) | 0.00 (1.53e-05 - 0.15) | 1* |
| SDF | 0.56 (0.09 - 1.79) | 0.43 (0.09 - 1.79) | 0.87 | 0.03 (1.41e-05 - 0.14) | 0.00 (1.41e-05 - 0.14) | 1* |
| overall | 0.59 (0.10 - 1.82) | 0.46 (0.10 - 1.82) | 0.86 | 0.03 (1.50e-05 - 0.15) | 0.00 (1.50e-05 - 0.15) | 1* |
| ***Pra-Offin*** |  |  |  |  |  |  |
| *SDF* | 0.63 (0.09 - 1.93) | 0.50 (0.09 - 1.93) | 0.83 | 0.03 (2.13e-05 - 0.18) | 0.01 (2.13e-05 - 0.18) | 1* |
| *RF* | 0.82 (0.08 - 3.04) | 0.57 (0.08 - 3.04) | 0.74 | 0.03 (2.14e-05 - 0.27) | 0.01 (2.14e-5 - 0.27) | 1* |
| *CS* | 0.72 (0.08 - 2.63) | 0.50 (0.78 -2.63) | 0.78 | 0.03 (2.05e-05 - 0.23) | 0.01 (2.05e-05 - 0.23) | 1* |
| *overall* | 0.72 (0.08 - 2.56) | 0.52 (0.08 - 2.56) | 0.78 | 0.03 (2.10e-05 - 0.22) | 0.01 (2.10e-05 - 0.22) | 1* |
| ***Tano-Ankobra*** |  |  |  |  |  |  |
| *TZ* | 1.46 (0.32 - 3.99) | 1.22 (0.32 - 3.99) | 0.38 | 0.35 (0.01 - 1.89) | 0.16 (0.01 - 1.89) | 0.94 |
| *SDF* | 1.38 (0.28 - 3.98) | 1.11 (0.28 - 3.98) | 0.44 | 0.33 (0.01 - 1.75) | 0.15 (0.01 - 1.75) | 0.95 |
| *RF* | 1.76 (0.24 - 6.28) | 1.28 (0.24 - 6.28) | 0.38 | 0.42 (0.01 - 2.41) | 0.17 (0.01 - 2.41) | 0.92 |
| *overall* | 1.53 (0.28 - 4.84) | 1.20 (0.28 - 4.84) | 0.4 | 0.37 (0.01 - 2.04) | 0.16 (0.01 - 2.04) | 0.94 |

### Predicted and projected risk maps across the national and district-level

Predicted prevalence in 2015 showed that while many areas in northern Ghana achieved near-zero mf prevalence, some areas, predominantly in the eastern, middle, and southern Ghana, are still well above the WHO-recommended elimination threshold of 1% skin mf prevalence (Figure 5). By 2030, more areas were predicted to fall below the elimination mark, with the main exceptions being nine districts located in eastern (Nkwanta South, Kadjebi, Nkwanta North, Krachi East) and western (Ahanta West, Sekondi Takoradi, Effia Kwesimintsim, Shama, Komenda-Edina-Eguafo-Abirem) Ghana (Figure.6). We also observed areas in southern Ghana including the Greater Accra Region, showing ∼10% probability of prevalence being above 1% by 2030 (Figure 6).

**Figure 5.**
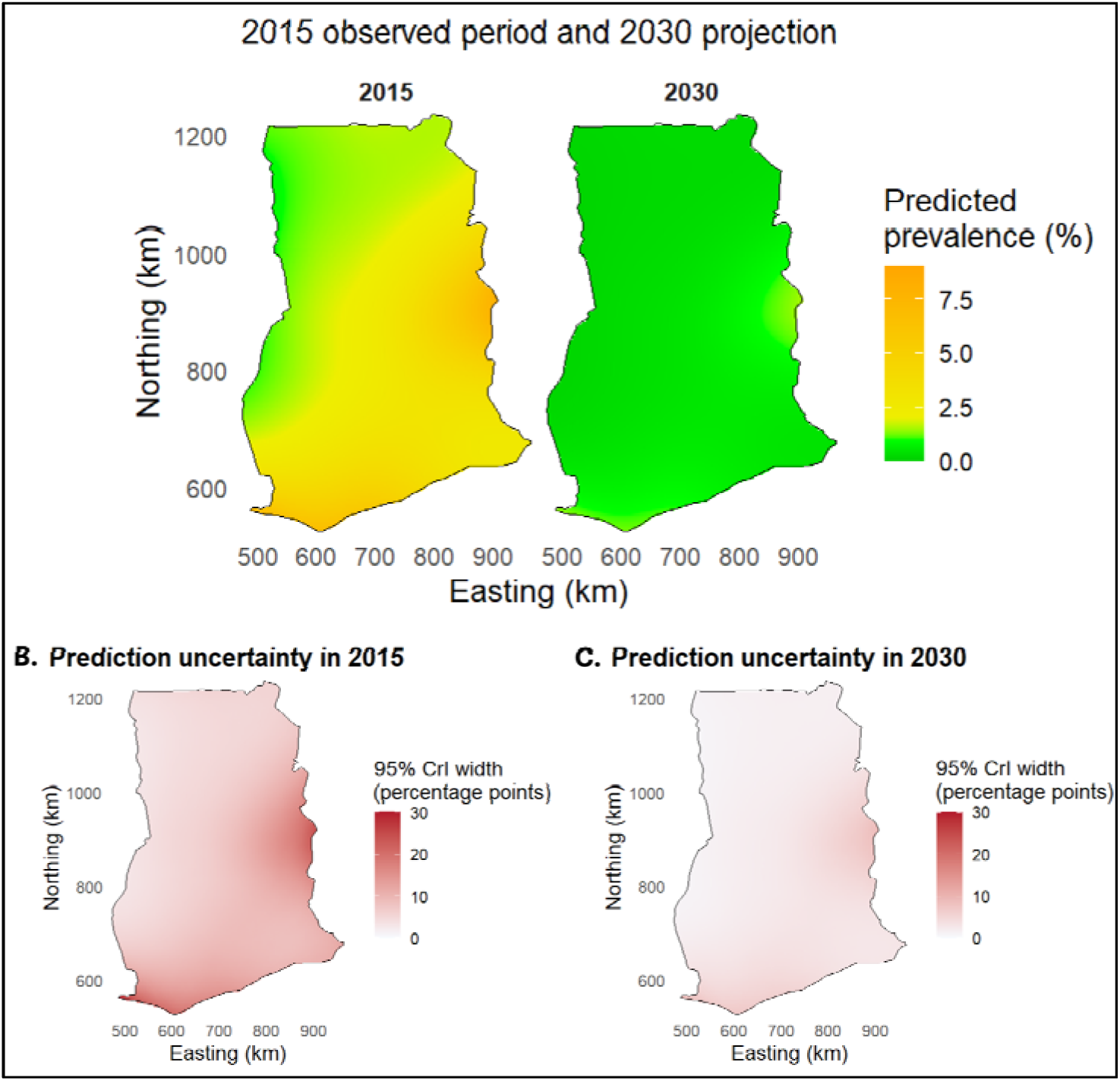
Posterior predicted prevalence in 2015 and 2030. Population-level 2D prediction (*t_2_*(easting, northing) on ∼2km grid. Infection risk varies, with the highest predicted infection in eastern and southwestern Ghana. Areas in eastern and the southwest show persistent hotspots.

**Figure 6.**
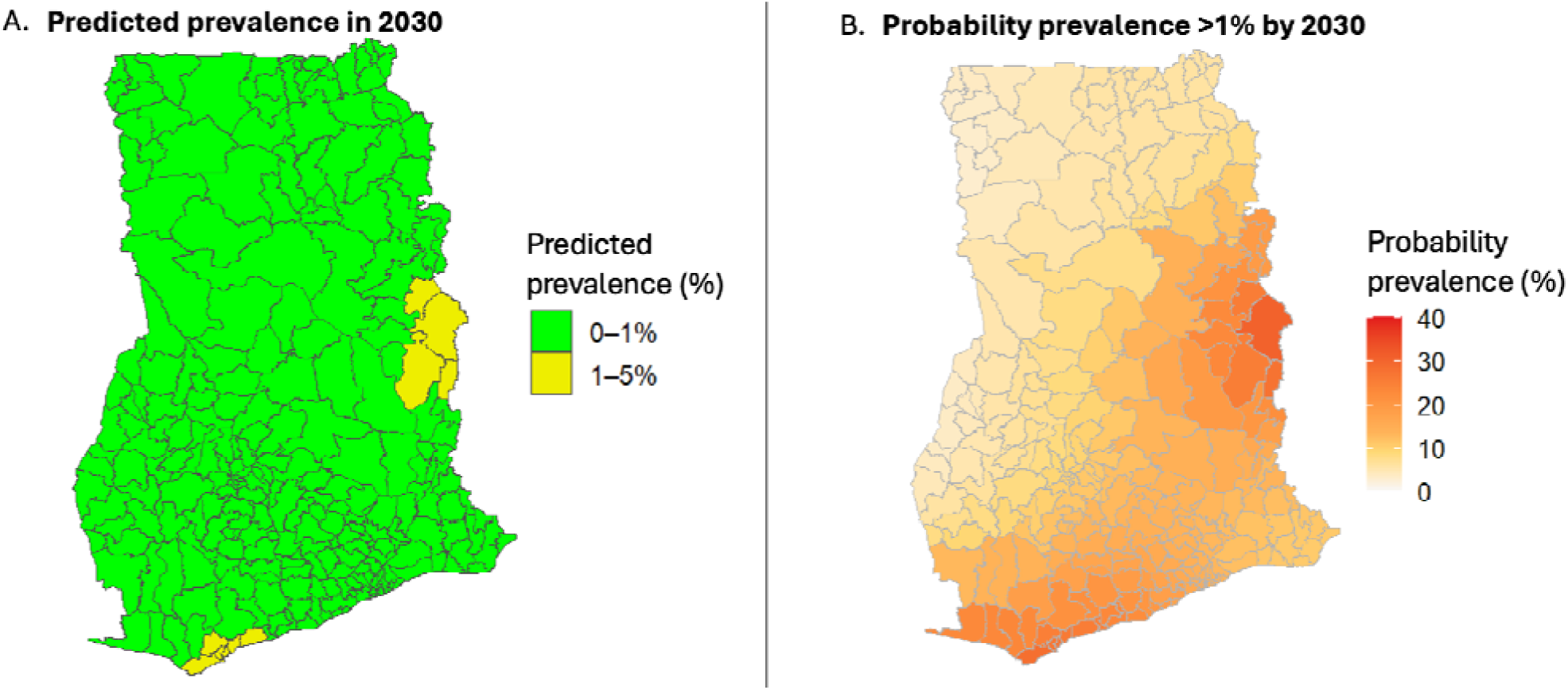
Predicted onchocerciasis prevalence. (A) District predicted prevalence in 2030. Yellow districts remain above 1%. (B) Probability that districts remain above 1%. Probabilities are higher (>10%) for districts in the eastern, southern and south-west of Ghana, shown in dark orange on map. This implies that there is a higher probability that these areas may not reach elimination by 2030.

## Discussion

Ghana has a long history of onchocerciasis, and, despite nearly a century of interventions, there are still persistent hotspots that pose a major challenge to the global elimination goal, warranting the need for tools that can enhance elimination efforts. While previous Ghana-based models of historical data have provided insights into historical trends, they did not incorporate spatial context or rigorous statistical methodologies and therefore lacked the power to make projections beyond the data (Biritwum et al., 2021). As such, this study presents the first comprehensive spatio-temporal analysis of onchocerciasis prevalence trends in Ghana from 1975 to 2015 and beyond. By integrating 1,353 village-level surveys spanning over four decades of programmatic intervention with climatic, socioeconomic, and environmental predictors, we have mapped onchocerciasis prevalence trends across Ghana’s operational transmission zones. While the overall results clearly show impressive progress toward elimination, we identified hotspots of persistent transmission where targeted interventions are needed.

### Drivers of onchocerciasis transmission and spatial heterogeneity

We hypothesised for drivers of onchocerciasis transmission. Our first driver, temperature, was strongly associated with longitude and hence was excluded from the final model to control for redundancy. Our second variable, which is hydrology, comprising surface wetness and runoff probability, showed a positive association with prevalence. Runoff probability, however, showed a non-linear plateau effect, indicating that beyond a threshold of hydrological suitability, additional runoff does not further increase transmission. This is biologically plausible: blackfly larval habitat is constrained by specific hydraulic conditions rather than simply increasing monotonically with water availability (Hougard et al., 1993; O’Hanlon et al., 2016). On the contrary, Shrestha et al. found no association between distance to the nearest river and onchocerciasis risk in Ghana’s Middle Transitional Zone. The authors suggested that this may have been due to the smaller geographic extent of their study area, where most locations were likely to be relatively close to a river, resulting in limited variation in distance to the nearest river and reducing its ability to discriminate differences in transmission risk. This contrasts with broader-scale modelling, such as studies covering Ethiopia, where substantial areas are unsuitable for onchocerciasis transmission and greater variation in distance to rivers may provide more informative spatial discrimination of risk. (Shrestha et al., 2024).

We had two hypothesised drivers associated with urbanisation: population density and land use. Contrary to our hypothesis, population density was not correlated with prevalence (Supp Figure 5). However, Land use variables associated with urbanisation were negatively associated with prevalence (Supp Figure 5). Urbanisation can decrease blackfly abundance through habitat modification, resulting in reduced exposure to vector breeding sites, whereas rural and less modified environments may sustain conditions favourable for vector persistence and transmission (Cheke, 2017; Nwoke et al., 2021). Urban environments can also decrease prevalence through improved access to health services and CDTI.

Prevalence was generally heterogeneous, characterised by a north-south spatial gradient captured by the 2D spatial smooth after controlling for all measured environmental predictors across Ghana. This variation in transmission intensity is known to be impacted by differences in blackfly species/subspecies composition and competence, which affect biting behaviour (Lamberton et al., 2014; Lamberton et al., 2015; Otabil et al., 2020). In addition, changes in the distribution of *Simulium damnosum* complex over time in response to environmental conditions further contribute to the differential transmission pattern (Post et al., 2013). Other unmeasured geographic factors impacting this gradient could include historical intervention intensity or landscape connectivity. The OCP activities were initially focused on high-endemic areas in northern and parts of central Ghana, while large areas of southern Ghana, particularly forested regions, were not included in the original vector-control programme (Biritwum et al., 2021; Lamberton et al., 2015). As a result, the historical geography of intervention was itself an important determinant of subsequent transmission patterns. The transition from aerial larviciding to ivermectin-based strategies, followed by the expansion of CDTI, produced major reductions in infection prevalence but also generated a landscape in which different areas experienced different durations, intensities and combinations of control (Biritwum et al., 2021; Kenneth Bentum Otabil et al., 2023). As such, areas that received early and sustained interventions may have experienced prolonged reductions in both parasite prevalence and vector abundance.

Spatial predictions also revealed a clear temporal transition in Ghana’s onchocerciasis burden consistent with the phased roll-out of control strategies. All OTZs started at high mean prevalence in 1975, with the steepest overall declines occurring between 1985 and 1999, coinciding with the expansion of OCP larviciding and the introduction of ivermectin-based treatment (APOC, 2003; Baker et al., 1990; World Health Organization, 2015). This reduction highlights the importance of consistent and sustained efforts to drive elimination (Nikièma et al., 2024; Ramirez, 2013). Projections to 2030 suggest near-elimination across most of Ghana, with all seven OTZs projected at or below the WHO 1% elimination threshold for mean predicted prevalence, although with wide credible intervals reflecting uncertainty beyond the observation window (Table 2). Moreover, these extrapolations, which we acknowledge require adequate caution in their interpretation, and assume that current levels of intervention remain.

### Probability of elimination and residual transmission risk

A recurring theme in these results is that the predicted probability of elimination of onchocerciasis in Ghana varies depending on the spatial scale at which it was predicted. Based on extrapolated prevalence rates from 2015, the White Volta-Kulpawn, Black Volta, Dayi-Asukawkaw and Tano-Ankobra OTZs do not have a 95% probability of reaching the 1% threshold by 2030, although they will be close (Table 2). However, for OTZs that span multiple ecozones, the risk of persistent transmission hotspots within those OTZs varied. In northern Ghana, both the Black and White Volta-Kulpawn OTZs traverse the Sudan and Guinea savannas. These areas are known to harbour the Savanna-adapted *Simulium* subspecies, which are substantially more anthropophilic than forest-adapted species, have higher vectorial capacity, and can thus maintain higher transmission levels within a population (Lamberton et al., 2014). Analysis at the ecozone level revealed differences between the two savanna ecozones: the Guinea savanna areas of the White Volta-Kulpawn OTZ had a lower probability (∼2%) of achieving a prevalence rate below 1% by 2030 relative to the Sudan savanna areas. Hence, it requires attention as it may potentially harbour transmission hotspots. Similarly, rainforest ecozone areas within the Tano-Ankobra OTZ appear less likely to attain 1% than semi-deciduous forest. This accords with recent mf prevalence surveys in western Ghana within the highly dense forest areas (Ahiadorme et al., 2026) and is consistent with higher transmission indices in forest *Simulium* subspecies (Cheke & Garms, 2013).

The extrapolated, fine-scale spatial risk maps reveal a ∼10% to 40% probability that predicted prevalence would be above 1% in areas of the Oti, Volta, and Western regions and along parts of the coast of southern Ghana (Figures 5 & 6). The Oti and Volta regions along the Ghana-Togo border (i.e., the Asukawkaw Ferry waterway) started larviciding in 1981 and were later included in the southeastern extension of the OCP larviciding intervention, and ivermectin intervention started in 1987. Historically, transmission has been high in the Oti-Volta area and was even considered a special intervention zone (SIZ), an area with >50% mf prevalence where larviciding continued after the OCP ended (Lamberton et al., 2015; Vinkeles Melchers et al., 2024). In contrast, southern Ghana was considered a low-transmission area, and no larviciding was done (Lamberton et al., 2015).

Similarly, Ahiadorme and colleagues (Ahiadorme et al., 2026) also reported an elevated probability of onchocerciasis infection in the far north (within the White Volta-Kulpawn), in the north-east and around Lake Volta, and in regions in the south along the Ghana-Côte d’Ivoire border (predominantly in the Eastern and Western regions, respectively). Although the estimates reported in our study are extrapolations and often close to the WHO elimination threshold, they nevertheless provide an objective indication of potentially persistent hotspots or residual transmission foci. Greater Accra is the only region considered non-endemic for onchocerciasis; however, with ∼10% probability of prevalence being >1% in 2030. In the last 3 years, there were reports of blackflies in the Oyarifa area. Recent (2023) epidemiological surveys in the La Nkwantanang district in the Greater Accra Region indicate transmission levels well above the elimination threshold. Mining activities (galamsey) have been reported on the Offin River and have been linked to the disappearance of blackflies in known breeding sites, as rivers are polluted and no longer suitable for the aquatic stages (Sumboh et al., 2026), and may contribute to their dispersal into areas originally considered unsuitable or non-endemic there is a need for mapping surveys to ascertain onchocerciasis transmission in the area.

Similarly, Ahiadorme et al. (2026) identified an elevated probability of onchocerciasis infection in northern Ghana, particularly within the White Volta–Kulpawn, in the north-east and around Lake Volta, as well as in parts of southern Ghana along the Ghana–Côte d’Ivoire border. Although these estimates are model-based extrapolations and, in some areas, approach the WHO elimination threshold, their spatial concordance with known ecological and epidemiological patterns provides an indication of areas where residual or persistent transmission may warrant further investigation. This is particularly relevant for Greater Accra, which is generally considered non-endemic for onchocerciasis, but where our model estimated an approximately 10% probability that prevalence could exceed 1% by 2030.

Over the past three years, blackflies have been reported in the La Nkwantanang–Madina area of the Greater Accra Region (Badu, 2023). Follow-up epidemiological surveys reportedly identified *O. volvulus* seropositivity levels above the WHO elimination threshold (unpublished data). The detection of blackflies alongside evidence suggestive of ongoing transmission challenges the assumption that transmission is absent from the region and highlights the need for periodic reassessment of areas historically classified as non-endemic, particularly where environmental and demographic conditions are changing.

At the same time, substantial environmental changes are occurring within river systems that have historically supported *Simulium* breeding. Recent work in the Offin River basin has documented extensive ecological degradation associated with illegal small-scale mining, popularly known in Ghana as “galamsey”, including increased sedimentation and turbidity, vegetation loss, and riparian disturbance (Sumboh et al., 2026). These changes may reduce the suitability of affected sites for the aquatic stages of blackflies and alter the distribution of suitable breeding habitats (Cheke, 2017; Lamberton et al., 2014; Sumboh et al., 2026; Wilson et al., 2002). Because parts of Greater Accra fall within the Pra–Offin operational transmission zone and are connected to broader riverine systems, environmental changes in neighbouring river basins could potentially influence the distribution, abundance, and dispersal of blackfly populations across the region. However, the extent to which environmental degradation has altered vector movement or contributed to the presence of blackflies in Greater Accra remains unknown. This uncertainty further highlights the need for targeted entomological and epidemiological surveillance to determine whether the detection of blackflies and *O. volvulus* exposure reflects local persistence, recent introduction, or previously unrecognised transmission.

Together, these findings highlight an important surveillance gap. Areas classified as non-endemic may not necessarily be epidemiologically static, while environmental changes may alter the distribution of suitable vector habitats and the connectivity of transmission systems. Given the capacity of members of the *S. damnosum* complex for long-distance Acdispersal (Cheke et al., 2024), and the potential for vector populations to move between connected ecological systems (Cheke et al., 2023; Cheke et al., 2017), targeted entomological and epidemiological surveys are needed to determine whether the elevated modelled risk in Greater Accra reflects residual transmission, localised transmission foci, or uncertainty arising from limited surveillance data. Such surveys would be particularly valuable around river systems and other areas where human populations may be exposed to dispersing blackflies.

### Implications for elimination programmes

Our findings confirm that Ghana had achieved near-elimination of onchocerciasis nationally by 2015, with median mf prevalence in most districts predicted to be below the WHO 1% elimination threshold. However, some areas still showed persistent and possible transmission emergence.

The spatial heterogeneity in residual transmission, with lower probabilities of achieving a <1% mf prevalence in the White Volta-Kulpawn and Tano-Ankobra, highlights that a uniform national approach might be insufficient to achieve the WHO 2030 elimination goal. Furthermore, the marginal probabilities of >1% mf prevalence in southern Ghana stress the need for mapping in historically non-endemic areas, and an urgent need for follow-up surveys to validate or refine model predictions of progress toward elimination. Our analysis reveals persistence of geographically coherent transmission foci in the White Volta-Kulpawn, Tano-Ankobra OTZ, eastern corridors (comprising the Nkwanta South, Kadjebi, Nkwanta North, and Krachi East districts along the Oti river basin and Asukawkaw ferry in the east), and southwestern coast (made of Ahanta West, Sekondi Takoradi, Effia Kwesimintsim, Shama, Komenda-Edina-Eguafo-Abirem districts within the Rain Ahanta West, Sekondi Takoradi, Effia Kwesimintsim, Shama, Komenda-Edina-Eguafo-Abirem districts within the Rain Forest ecozone). In particular, we recommend surveys targeting areas in the Greater Accra Region and regions within the rain forest ecozones, preferably with sensitive tools such as PCR from skin snips where feasible or OV16 serological tests to detect exposure to the parasite.

### Model performance and limitations

The final model showed excellent convergence and good predictive performance across the full prevalence range, and the model accounted for almost all of the variation in prevalence rates (R^2^ = 0.897, p<0.001) (Supp Figure 3). We observed slight underprediction at high observed prevalence values consistent with partial pooling toward the population mean in the hierarchical model structure. This shrinkage is expected and well-characterised in Bayesian hierarchical models (Gelman & Hill, 2006) and does not compromise inference for the primary study period.

However, some limitations and caveats are worth highlighting. Firstly, approximately 0.3% of observations had high Pareto k (k >1.0) values identified from LOO-CV, indicating influential observations (Vehtari et al., 2024). These were concentrated in high-prevalence, early surveys and survey clusters, a known challenge in hierarchical models fitted to spatially clustered epidemiological data (Adin et al., 2024; Roberts et al., 2017). Secondly, the extrapolation of temporal smooths beyond the observation period (1975 - 2015) assumes that trends continue as fitted, which may not hold if intervention coverage, vector ecology, or climate change occur in ways not captured by the model. Thirdly, the cross-sectional survey design precludes causal inference on the effects of specific interventions, as temporal and spatial confounding between intervention phases and other predictors cannot be fully disentangled.

Here, the buffered testing clinic locations were used as the point estimate for prevalence within the community. The results would be significantly improved if future data collection included a metadata record of the participant’s home location, i.e., allowing records of village locals to be differentiated from visitors and enabling correlation between actual local conditions and infection rate.

Finally, this study relied on skin-snip microscopy to estimate *O. volvulus* microfilarial (mf) prevalence. Although microscopy is useful for detecting infection in high-endemic settings, its sensitivity decreases as parasite burden declines, meaning that prevalence may have been underestimated in areas with prolonged community-directed treatment with ivermectin (CDTI). Conversely, microscopy may also have overestimated *O. volvulus* prevalence because *Mansonella streptocerca* mf occur in skin snips and must be differentiated from *O. volvulus* on the basis of morphology, which may be challenging in routine field-based microscopy (Mathison et al., 2019). Recent work in Ghana (unpublished) detected *M. streptocerca* sequences in skin-snip samples that were initially positive for *O. volvulus* by microscopy but subsequently tested negative by PCR, highlighting the potential for species misclassification in microscopy-based surveys. Future studies should therefore incorporate more sensitive and species-specific diagnostic approaches, including PCR and antigen-based assays, which may improve detection at low infection burdens and distinguish *O. volvulus* from morphologically similar filarial species (Fischer et al., 1998).

## Conclusion

Spatio-temporal modelling of existing surveillance data provides a cost-effective and statistically rigorous approach to prioritising mapping efforts and can be relevant in the onchocerciasis context (Ogunmiloro & Idowu, 2024; Shrestha et al., 2022). The ∼2 km grid predictions generated by this model provide fine-scale, spatially explicit risk estimates that can directly inform decisions about where to deploy new mapping surveys, where to intensify or maintain CDTI, and where stop-MDA decisions could be considered (WHO, 2025). This study presents the first comprehensive Bayesian spatio-temporal analysis of onchocerciasis mf prevalence across Ghana, integrating 1,353 village-level surveys spanning four decades of programmatic intervention. Our findings confirm that Ghana had achieved near-elimination of onchocerciasis by 2015, with substantial declines across all seven OTZs, six agro-ecological zones, and 147 districts following the sequential roll-out of aerial larviciding, ivermectin mobile teams, and annual and biannual CDTI. Most transmission zones were projected below the WHO 1% elimination threshold, and most districts that carried the highest burden are expected to have reached or fall below the elimination threshold by 2030.

These findings provide a spatially explicit evidence-based prioritisation of resources and interventions in Ghana’s onchocerciasis elimination programme. Persistent transmission foci in southern Ghana, particularly in forest zones, eastern and southwestern corridors and districts as well as communities near the Togo border, represent areas to prioritise for targeted investigation and intensified intervention to meet WHO elimination thresholds.

## Data Availability

All data are available online at the ESPEN portal.

https://espen.afro.who.int/countries/ghana

## Acknowledgements

The authors wish to thank Warwick Grant, Sindew Feleke, and Bright Asare at La Trobe University (Australia), Gabriella Ondoua Nganjou at the University of Yaoundé, and Annette Kuesel at UNICEF/UNDP/World Bank/WHO Special Programme for Research and Training in Tropical Diseases for helpful discussions on onchocerciasis in Africa, and Samuel Oppong at The KIDS Research Institute Australia for discussions around modelling frameworks.

## Author Contributions

Conceptualization: MO, SMH, KKF, DAB; Data Curation: MO, KKF, DdS, SP-B, JHNO, JO, EM, ODA; Formal analysis: MO, DD; Funding acquisition: SMH, KKF, DAB; Investigation: MO; Methodology: MO, DD, HS; Project administration: SMH, KKF, DAB; Supervision: SMH, KKF, DAB; Validation: DD, SMH; Writing, original draft: MO; Writing, review & editing: MO, SMH, DD, DdS, SP-B, JHNO.

