## Supplemental for "A geospatial approach to understanding transmission patterns of onchocerciasis in Ghana"

This Supplemental document provides additional information organised as follows.

- Data sources (Supp Table 1, 2)
- Predictor pre-processing (Supp Table 3, Figure 1)
- Model performance and diagnostics (Supp Table 4; Supp Figure 2-3).
- Posterior predictions (Supp. Table 5, Supp. Figure 4- 5)

### Data sources

Supp Table 1: Bioclimatic and environmental variables

| # | Variables | Description | Source | Resolution | Scale | Duration |
| --- | --- | --- | --- | --- | --- | --- |
| 1 | Landcover | MCD12Q1.061 MODIS Land Cover Type Yearly Global 500m | [NASA LP DAAC at the USGS EROS Center](https://doi.org/10.5067/MODIS/MCD12Q1.061) |  | 1000 | 2001-2015 |
| 2 | Flow accumulation | WWF HydroSHEDS Flow Accumulation | https://www.hydrosheds.org/ |  | 30 Arc-Seconds | 2000 |
| 3 | Flow accumulation | NASA | [NASA LP DAAC at the USGS EROS Center](https://doi.org/10.5067/VIIRS/VNP21A1D.002), https://doi.org/10.5067/VIIRS/VNP21A1D.002 |  |  | 2012-2015 |
| 4 | Land surface Temperature | MODIS/006/MOD11A1 | MODIS/006/MOD11A1 |  | 1000 | 1975-2015 |
| 5 | Population density | CIESIN/GPWv411/GPW_UNWPP-Adjusted_Population_Density | https://doi.org/10.7927/H4F47M65 |  |  | 2015 |
| 10 | Slope | [NASA/CGIAR](https://srtm.csi.cgiar.org/) | [SRTM Digital Elevation Data Version 4  \|  Earth Engine Data Catalog  \|  Google for Developers](https://developers.google.com/earth-engine/datasets/catalog/CGIAR_SRTM90_V4#description)  Estimated from elevation | 90 meters | 1000 | 2000 |
| 12 | Elevation | [NASA/CGIAR](https://srtm.csi.cgiar.org/) | [SRTM Digital Elevation Data Version 4  \|  Earth Engine Data Catalog  \|  Google for Developers](https://developers.google.com/earth-engine/datasets/catalog/CGIAR_SRTM90_V4#description) | 90 meters | 1000 | 2000 |
| 15 | NDVI_mean |  |  |  |  |  |
| 16 | EVI_mean |  |  |  |  |  |
| 17 | Soil_moist |  |  |  |  |  |
| 18 | TCW_wetness | Malaria Atlas Project | projects/malariaatlasproject/assets/TCW_v061/1km/Monthly | 5000 m |  | 2001-2015 |
| 19 | Temp_avg | MODIS/Terra Land Surface Temperature/Emissivity  Daily L3 Global 1km SIN Grid V061 | [MODIS/Terra Land Surface Temperature/Emissivity Daily L3 Global 1km SIN Grid V061 \| NASA Earthdata](https://www.earthdata.nasa.gov/data/catalog/lpcloud-mod11a1-061) | LST_Day_1km | 30 degree  1000x1000 m | 2000-2015 |
| 20 | Chirps_precip | [CHIRPS: Rainfall Estimates from Rain Gauge and Satellite Observations \| Climate Hazards Center - UC Santa Barbara](https://chc.ucsb.edu/data/chirps) | UCSB-CHG/CHIRPS/DAILY | 1000 | 0.05°  1000 | 1981-2015 |
| 21 | Rural_Urban | JRC/GHSL/P2023A/GHS_SMOD_V2-0/2030 Population count by epoch | [GHSL: Global population surfaces 1975-2030 (P2023A)  \|  Earth Engine Data Catalog  \|  Google for Developers](https://developers.google.com/earth-engine/datasets/catalog/JRC_GHSL_P2023A_GHS_POP) | 100 m | 1000 | 1975-2015 |

Supp Table 2: Bioclimatic variables

| # | Bioclimatic variable | Predictors interpretation |
| --- | --- | --- |
| 1 | bio1 | Annual mean temp |
| 2 | bio2 | Mean daily range (Mean of monthly (max temp - min temp)) |
| 3 | bio3 | Isothermality (BIO2/BIO7) (×100) |
| 4 | bio4 | Temperature Seasonality (standard deviation ×100) |
| 5 | bio5 | Max Temperature of Warmest Month |
| 6 | bio6 | Min Temperature of Coldest Month |
| 7 | bio7 | Average temperature range |
| 8 | bio8 | Mean temp of wettest quarter |
| 9 | bio9 | Mean temp of driest quarter |
| 10 | bio12 | Annual precipitation |
| 11 | bio13 | Precipitation of wettest month |
| 12 | bio14 | Precipitation of driest month |
| 13 | bio15 | Rainfall seasonality (CV) |
| 14 | bio16 | Precipitation of wettest quarter |
| 15 | bio17 | Precipitation of driest quarter |
| 16 | bio18 | Precipitation of warmest quarter |
| 17 | bio19 | Precipitation of warmest quarter |

Supp Table 3: Variable Selection and Model Development

Results of forward selection using R package adespatial, function forward_sel() to identify variables to be retained in spatial model.

| variables | order | R^2^ | R^2^Cum | AdjR^2^Cum | F | pvalue |
| --- | --- | --- | --- | --- | --- | --- |
| bio9 | 27 | 0.020 | 0.020 | 0.020 | 28.88 | 0.001 |
| bio16 | 34 | 0.022965206 | 0.04368525 | 0.04228097 | 32.707443 | 0.001 |
| slope | 16 | 0.025804070 | 0.06948932 | 0.06743823 | 37.742006 | 0.001 |
| bio8 | 26 | 0.010257878 | 0.07974719 | 0.07704057 | 15.159654 | 0.001 |
| Bio13 | 31 | 0.011321497 | 0.09106869 | 0.08772457 | 16.927477 | 0.001 |
| waterway_length | 38 | 0.003505454 | 0.09457414 | 0.09057374 | 5.257643 | 0.017 |
| bio18 | 36 | 0.004222453 | 0.09879660 | 0.09414780 | 6.358020 | 0.012 |
| bio3 | 21 | 0.006875101 | 0.10567170 | 0.10039542 | 10.424177 | 0.004 |
|  |  |  | p < 0.05 &> |  |  |  |
| elevation | 17 | 0.002380715 | 0.10805241 | 0.10212804 | 3.616657 | 0.054 |
| temp_avg | 3 | 0.003803156 | 0.11185557 | 0.10529616 | 5.798013 | 0.026 |
| Land_cover | 6 | 0.003501082 | 0.11535665 | 0.10816443 | 5.354660 | 0.021 |
| TCW_wetness | 8 | 0.001882070 | 0.11723872 | 0.10940356 | 2.882499 | 0.082 |
| Flow_accumulation | 5 | 0.002319268 | 0.11955799 | 0.11108593 | 3.558817 | 0.076 |
| bio17 | 35 | 0.001291465 | 0.12084945 | 0.11173234 | 1.983139 | 0.155 |

### Supp 2.3 Predictor pre-processing

All potential predictors were first assessed for cross-correlation, with many having correlations exceeding 0.9.

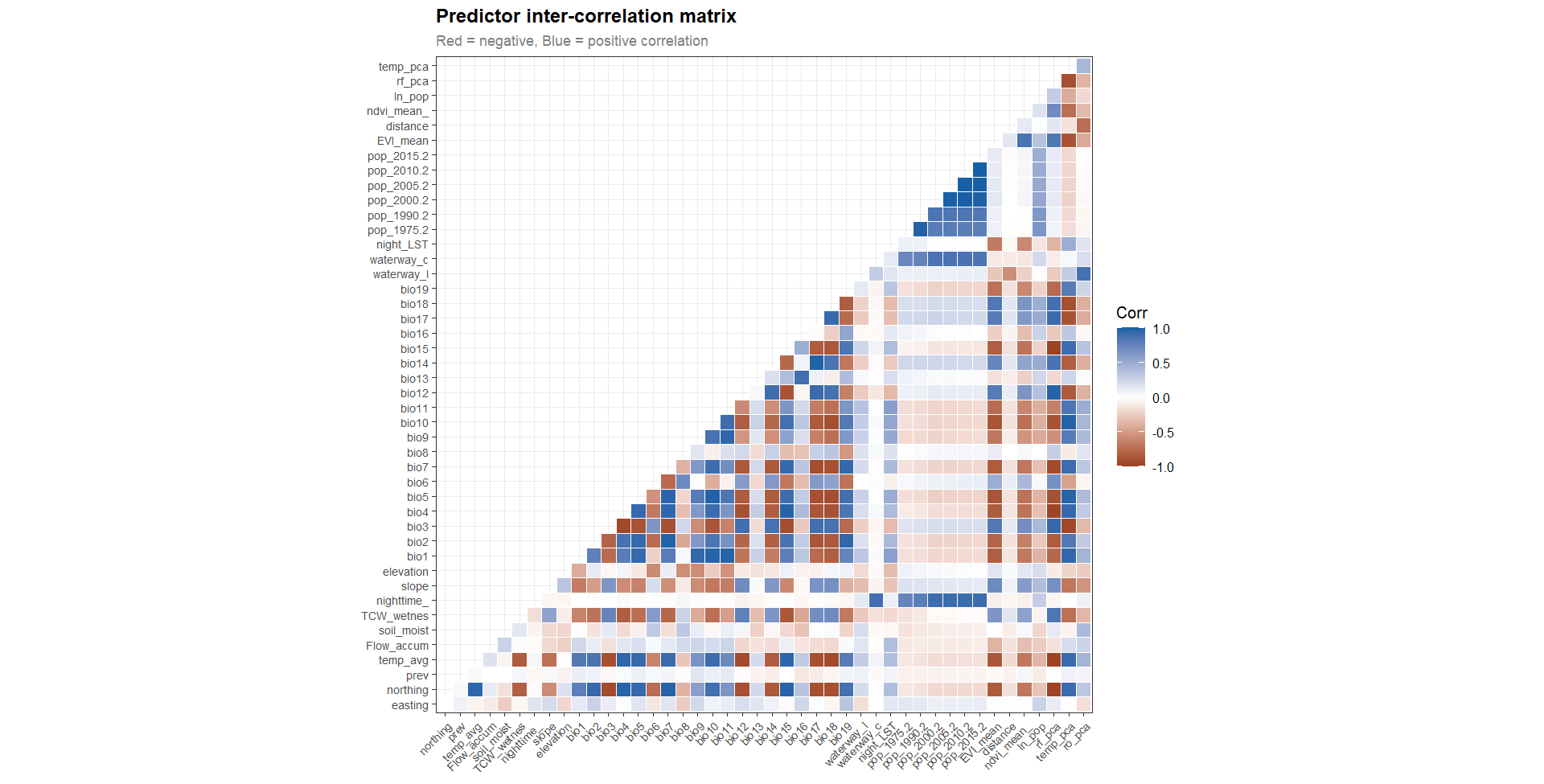

Supp Figure 1: Correlation between continuous variables. Where dark blue and red indicate strong positive and negative correlation.

#### Supp 2. 4 Composite predictor

Because of the high correlation between multiple predictors and to maximise model parsimony and interpretation, we used principal components analysis to extract the maximum variation from combinations of predictors to capture the spatial phenomena corresponding to each hypothesis in the time series model of prevalence.

Calculation of each composite variable followed the same workflow, where those variables considered likely to represent the hypotheses best were first selected using forward selection based on their correlation with prevalence. Selected predictors were standardized to zero mean and unit variance. A principal component analysis was then run on the standardized predictors and the first PCA was extracted for use as the composite variable.

**For temperature (bio1, bio3)**

tempdat <- data.frame(tave = x$bio1, tcv = x$bio3)

temp.std <- apply(tempdat, 2, scale)

temp_pca <- princomp(temp.std)

temp_pca$loadings

\# add to df

moddat$temp_pca <- temp_pca$scores[,1]

**runoff probability composite (PCA)**

rocof <- data.frame(wway_length = x$waterway_length,

dist = x$distance,

ln_slope = log(x$slope),

soil_moist = x$soil_moist)

rocof.std <- apply(rocof, 2, scale)

cor(rocof.std, x$prev)

ro_pca <- princomp(rocof.std)

summary(ro_pca)

ro_pca$loadings

\# add composite rainfall and runoff prob

moddat$ro_pca <- ro_pca$scores[,1] # 45% of variance

**Population density PCA**

ln_pop <- log(numeric_cols[,48:53]) # 48 to 53 contain the pop density values for 1975-2015

ln_pop <- apply(ln_pop, 2, scale)

pop.pca <- princomp(ln_pop) # principal component analysis

moddat$ln_pop <- pop.pca$scores[,1]

### Supp 2 Table 4: Model diagnostics: Data records with pareto-k values >1

| village | year | Positive | Examined | OTZ | ecozone | Prevalence (%) |
| --- | --- | --- | --- | --- | --- | --- |
| Awate Todzi (Agadome) | 1989 | 8 | 114 | Dayi-Asukawkaw | TZ | 7 |
| Biakpa | 1978 | 105 | 389 | Dayi-Asukawkaw | SDF | 27 |
| Okanease | 1990 | 128 | 171 | Oti-Daka | TZ | 74.9 |
| Kpasera | 1989 | 3 | 210 | Pru-Afram | GS | 1.4 |

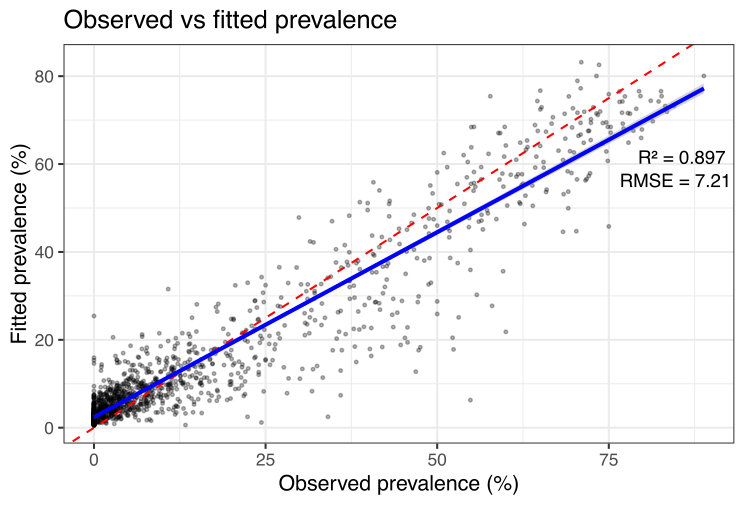

Supp Figure 2. Observed versus fitted microfilarial prevalence. Scatter plot of posterior mean fitted values (x-axis) against observed prevalence (y-axis) for all 1,353 survey observations. The red loess smoother closely follows the dashed 1:1 identity line across most of the prevalence range (Pearson’s r = 0.947, R^2^ = 0.897, Root mean square error (RMSE) = 0.072, average absolute prediction error (MAE) = 0.0491, p < 0.001), indicating good model calibration. Slight underprediction at near-zero and very high prevalence values is consistent with partial pooling toward the population mean in the hierarchical model structure. This reflects the shrinkage of historically high pre-intervention estimates. The dense cluster of points near the origin reflects the high proportion of near-zero prevalence observations in the post-intervention period.

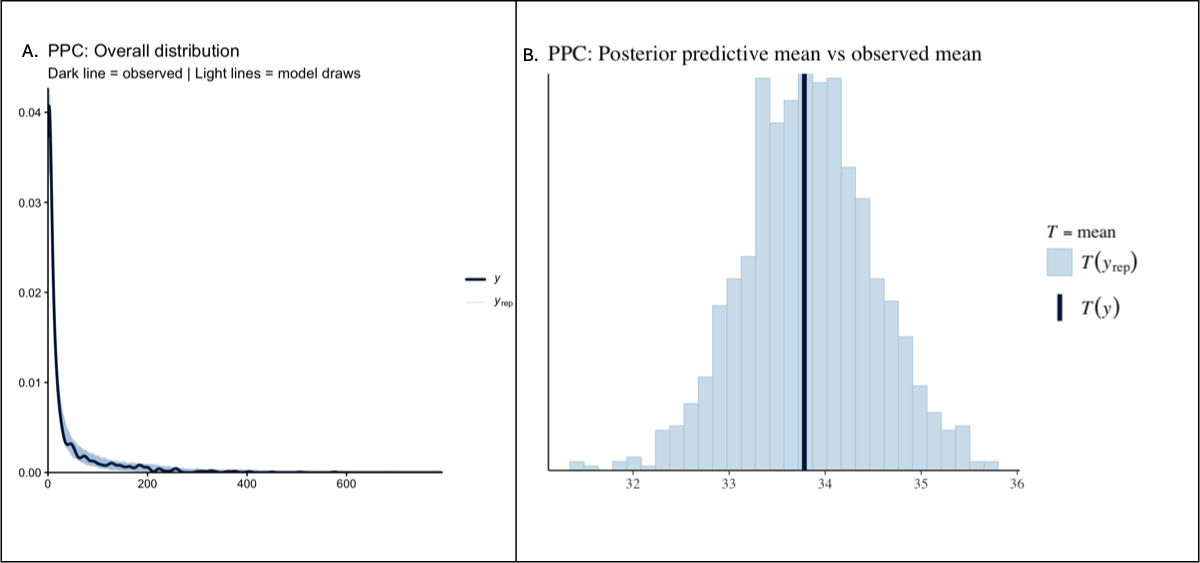

Figure 3. Posterior probability check plot. Panel A shows the overall distribution with observed data falling well within the model draws (1000 draws). Panel shows the posterior predictive mean versus the observed mean prevalence. The distribution appears to follow a bell-shape.

### S7. Posterior predictions

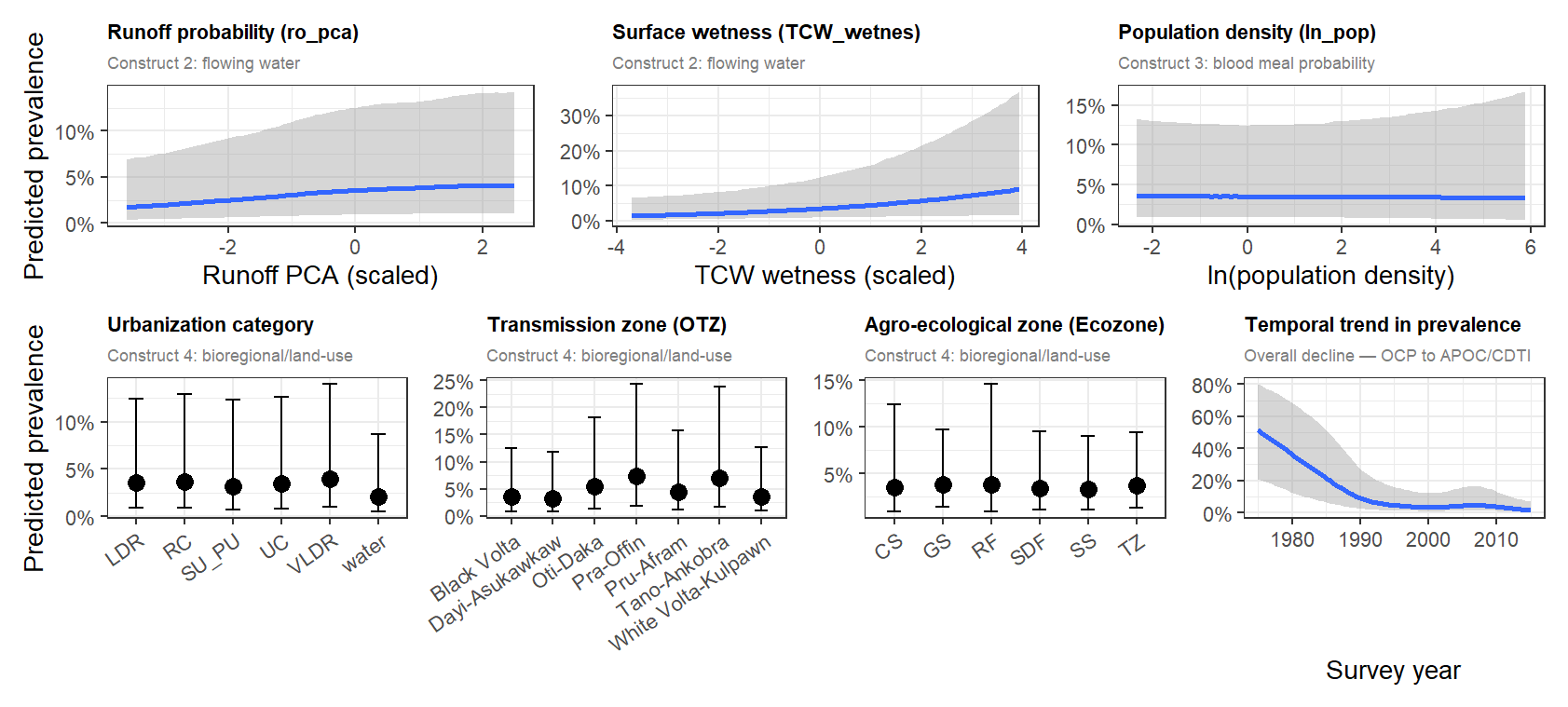

G. **posterior marginal effect plots** from your Bayesian zero-inflated beta-binomial model. They show how each predictor affects **predicted onchocerciasis prevalence**, while all other variables are held constant (typically at their mean or reference level). **posterior marginal effect plots** from your Bayesian zero-inflated beta-binomial model. They show how each predictor affects **predicted onchocerciasis prevalence**, while all other variables are held constant (typically at their mean or reference level). **posterior marginal effect plots** from your Bayesian zero-inflated beta-binomial model. They show how each predictor affects **predicted onchocerciasis prevalence**, while all other variables are held constant (typically at their mean or reference level).

F. **posterior marginal effect plots** from your Bayesian zero-inflated beta-binomial model. They show how each predictor affects **predicted onchocerciasis prevalence**, while all other variables are held constant (typically at their mean or reference level). **posterior marginal effect plots** from your Bayesian zero-inflated beta-binomial model. They show how each predictor affects **predicted onchocerciasis prevalence**, while all other variables are held constant (typically at their mean or reference level). **posterior marginal effect plots** from your Bayesian zero-inflated beta-binomial model. They show how each predictor affects **predicted onchocerciasis prevalence**, while all other variables are held constant (typically at their mean or reference level).

E. **posterior marginal effect plots** from your Bayesian zero-inflated beta-binomial model. They show how each predictor affects **predicted onchocerciasis prevalence**, while all other variables are held constant (typically at their mean or reference level). **posterior marginal effect plots** from your Bayesian zero-inflated beta-binomial model. They show how each predictor affects **predicted onchocerciasis prevalence**, while all other variables are held constant (typically at their mean or reference level). **posterior marginal effect plots** from your Bayesian zero-inflated beta-binomial model. They show how each predictor affects **predicted onchocerciasis prevalence**, while all other variables are held constant (typically at their mean or reference level).

D. **posterior marginal effect plots** from your Bayesian zero-inflated beta-binomial model. They show how each predictor affects **predicted onchocerciasis prevalence**, while all other variables are held constant (typically at their mean or reference level). **posterior marginal effect plots** from your Bayesian zero-inflated beta-binomial model. They show how each predictor affects **predicted onchocerciasis prevalence**, while all other variables are held constant (typically at their mean or reference level). **posterior marginal effect plots** from your Bayesian zero-inflated beta-binomial model. They show how each predictor affects **predicted onchocerciasis prevalence**, while all other variables are held constant (typically at their mean or reference level).

C. **posterior marginal effect plots** from your Bayesian zero-inflated beta-binomial model. They show how each predictor affects **predicted onchocerciasis prevalence**, while all other variables are held constant (typically at their mean or reference level). **posterior marginal effect plots** from your Bayesian zero-inflated beta-binomial model. They show how each predictor affects **predicted onchocerciasis prevalence**, while all other variables are held constant (typically at their mean or reference level). **posterior marginal effect plots** from your Bayesian zero-inflated beta-binomial model. They show how each predictor affects **predicted onchocerciasis prevalence**, while all other variables are held constant (typically at their mean or reference level).

B. **posterior marginal effect plots** from your Bayesian zero-inflated beta-binomial model. They show how each predictor affects **predicted onchocerciasis prevalence**, while all other variables are held constant (typically at their mean or reference level). **posterior marginal effect plots** from your Bayesian zero-inflated beta-binomial model. They show how each predictor affects **predicted onchocerciasis prevalence**, while all other variables are held constant (typically at their mean or reference level). **posterior marginal effect plots** from your Bayesian zero-inflated beta-binomial model. They show how each predictor affects **predicted onchocerciasis prevalence**, while all other variables are held constant (typically at their mean or reference level).

A. **posterior marginal effect plots** from your Bayesian zero-inflated beta-binomial model. They show how each predictor affects **predicted onchocerciasis prevalence**, while all other variables are held constant (typically at their mean or reference level). **posterior marginal effect plots** from your Bayesian zero-inflated beta-binomial model. They show how each predictor affects **predicted onchocerciasis prevalence**, while all other variables are held constant (typically at their mean or reference level). **posterior marginal effect plots** from your Bayesian zero-inflated beta-binomial model. They show how each predictor affects **predicted onchocerciasis prevalence**, while all other variables are held constant (typically at their mean or reference level).

**Figure 4. Posterior marginal effect plots** from the Bayesian zero-inflated beta-binomial model. These show how each predictor affects the **onchocerciasis predicted prevalence**, while all other variables are held constant. (A) Runoff probability (B) Surface wetness (C) Population density (D) Urbanization category (E) OTZ = Operational transmission zone (G) Temporal trend in prevalence. The blue and black dots represent the posterior mean predicted prevalence. Grey shaded region/error bars = 95% credible intervals (uncertainty), X-axis = predictor values or categories and Y-axis = predicted prevalence (%).

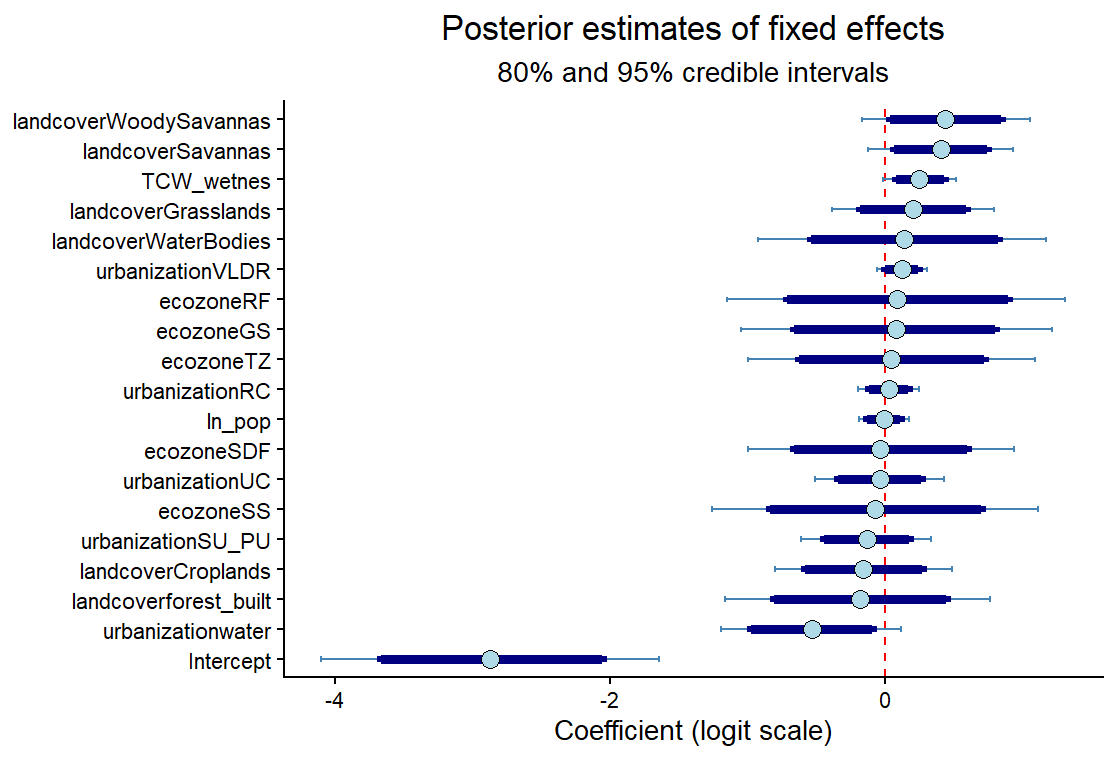

Figure 5. **Posterior distributions for model coefficients**. The Bayesian coefficient (forest) plot showing the posterior estimates of the model's fixed effects on the logit scale. The blue circle is the posterior mean estimate (the most likely coefficient value), the dark, thick blue bar represents 80% credible interval, the thin blue light bar indicates the 95% credible interval, and the red vertical dashed line at 0 indicates no effect. Bars crossing the red dashed lines indicate substantial uncertainty in their effects after accounting for other variables. The factor levels for landcover, ecozone and urbanization were Cropland/Natural Vegetation Mosaics, Coastal Savanna (CS) and low-density rural (LDR) reference

Table 5: Posterior probabilities of achieving the World Health Organization 1% onchocerciasis elimination threshold by 2015 and 2030 for Operational Transmission Zones (OTZ) within ecozones. * probability >95% of achieving elimination.

| **Ecozone** | | | | | | |
| --- | --- | --- | --- | --- | --- | --- |
|  | **2015** | | | **2030** | | |
|  | Mean (95% CI) | Median (95% CI) | Probability <1% | Mean (95% CI) | Median (95% CI) | Probability <1% |
| OTZ | Sudan Savanna | | | | | |
| Black Volta | 1.92 (0.48 - 4.94) | 1.63 (0.48 - 4.94) | 0.21 | 0.33 (0.01 - 1.80) | 0.13 (0.01 - 1.80) | 0.93 |
| White Volta-Kulpawn | 1.48 (0.37 - 3.77) | 1.27 (0.37 - 3.77) | 0.35 | 0.42 (0.01 - 2.30) | 0.19 (0.01 - 2.30) | 0.91 |
| Oti-Daka | 1.06 (0.28 - 2.65) | 0.92 (0.28 - 2.65) | 0.56 | 0.21 (0.01 - 0.87) | 0.13 (0.01 - 0.87) | 0.98* |
| overall | 1.48 (0.34 - 4.15) | 1.23 (0.34 - 4.15) | 0.37 | 0.32 (0.01 - 1.68) | 0.15 (0.01 - 1.68) | 0.94 |
|  | Guinea Savanna | | | | | |
| Black Volta | 2.20 (0.55 - 5.97) | 1.97 (0.55 - 5.97) | 0.13 | 0.40 (0.01 - 2.30) | 0.16 (0.01 - 2.30) | 0.9 |
| White Volta-Kulpawn | 1.76 (0.44 - 4.44) | 1.53 (0.44 - 4.44) | 0.24 | 0.50 (0.01 - 2.78) | 0.23 (0.01 - 2.78) | 0.88 |
| Oti-Daka | 1.26 (0.32 - 3.12) | 1.11 (0.32 - 3.12) | 0.43 | 0.25 (0.02 - 1.04) | 0.16 (0.02 - 1.04) | 0.97* |
| Pru-Afram | 0.58 (0.11 - 1.68) | 0.47 (0.11 - 1.68) | 0.87 | 0.03 (1.60e-05 - 0.14) | 0.00 (1.60e-05 - 0.14) | 1* |
| overall | 1.48 (0.18 - 4.50) | 1.19 (0.18 - 4.50) | 0.42 | 0.29 (0.00 - 1.81) | 0.11 (0.00 - 1.81) | 0.94 |
|  | Transitional Zone | | | | | |
| Black Volta | 2.24 (0.44 - 6.10) | 1.88 (0.44 - 6.10) | 0.18 | 0.39 (0.01 - 2.21) | 0.15 (0.01 - 2.21) | 0.91 |
| Oti-Daka | 1.25 (0.26 - 3.31) | 1.04 (0.26 - 3.31) | 0.47 | 0.24 (0.01 - 1.03) | 0.16 (0.01 - 1.03) | 0.97* |
| Dayi-Asukawkaw | 0.70 (0.10 - 2. 43) | 0.52 (0.10 - 2. 43) | 0.8 | 0.29 (0.00 - 2.54) | 0.03 (0.00 - 2.54) | 0.94 |
| Pru-Afram | 0.57 (0.09 - 1.75) | 0.45 (0.09 - 1.75) | 0.87 | 0.04 (1.48e-05 - 0.14) | 0.00 (1.48e-05 - 0.14) | 1* |
| Tano-Ankobra | 1.36 (0.26 - 3.86) | 1.11 (0.26 - 3.86) | 0.44 | 0.33 (0.01 - 1.73) | 0.15 (0.01 - 1.73) | 0.95 |
| overall | 1.22 (0.13 - 4.23) | 0.89 (0.13 - 4.23) | 0.55 | 0.26 (0.00 - 1.54) | 0.08 (0.00 - 1.54) | 0.95* |
|  | Semi-Deciduous Forest | | | | | |
| Dayi-Asukawkaw | 0.74 (0.11 - 2.64) | 0.54 (0.11 - 2.64) | 0.78 | 0.33 (0.00 - 2.85) | 0.03 (0.00 - 2.85) | 0.93 |
| Pru-Afram | 0.60 (0.10 - 1.83) | 0.47 (0.10 - 1.83) | 0.86 | 0.04 (1.70e-05 - 0.14) | 0.00 (1.70e-05 - 0.14) | 1* |
| Pra-Offin | 0.63 (0.10 - 1.86) | 0.50 (0.10 - 1.86) | 0.83 | 0.03 (2.17e-05 - 0.18) | 0.01 (2.17e-05 - 0.18) | 1* |
| Tano-Ankobra | 1.43 (0.28 - 4.03) | 1.17 (0.28 - 4.03) | 0.4 | 0.34 (0.01 - 1.81) | 0.16 (0.01 - 1.81) | 0.94 |
| overall | 0.85 (0.11 - 2.92) | 0.62 (0.11 - 2.92) | 0.79 | 0.18 (4.22e-05 - 1.21 | 0.02 (4.22e-05 - 1.21) | 0.97* |
|  | Coastal Savanna | | | | | |
| Dayi-Asukawkaw | 0.78 (0.09 - 3.04) | 0.51 (0.09 - 3.04) | 0.77 | 0.38 (0.00 - 3.01) | 0.03 (0.00 - 3.01) | 0.93 |
| Pra-Offin | 0.67 (0.07 - 2.41) | 0.47 (0.07 - 2.41) | 0.81 | 0.03 (1.93e-05 - 0.21) | 0.01 (1.93e-05 - 0.21) | 1* |
| overall | 0.72 (0.08 - 2.72) | 0.50 (0.80 - 2.72) | 0.79 | 0.20 (4.79e-05 - 1.41) | 0.01 (4.79e-05 - 1.41) | 0.97* |
|  | Rain Forest | | | | | |
| Pra-Offin | 1.01 (0.14 - 3.35) | 0.73 (0.14 -3.35) | 0.65 | 0.04 (3.00e-05 - 0.35) | 0.01 (3.0e-05 - 0.35) | 1* |
| Tano-Ankobra | 2.27 (0.42 - 7.45) | 1.77 (0.42 -7.45) | 0.22 | 0.54 (0.02 - 2.86) | 0.24 (0.01 - 2.86) | 0.89 |
| overall | 1.16 (0.18 - 5.95) | 1.18 (0.18 - 5.95) | 0.43 | 0.29 (8.34e-05 - 1.87) | 0.07 (8.34e-05 - 1.87) | 0.94 |
